# Loss of nucleoporin Nup50 is a risk factor for amyotrophic lateral sclerosis

**DOI:** 10.1101/2021.08.23.21262299

**Authors:** Salim Megat, Natalia Mora, Jason Sanogo, Alberto Catanese, Najwa Ouali Alami, Axel Freischmidt, Xhuljana Mingaj, Hortense de Calbiac, François Muratet, Sylvie Dirrig-Grosch, Stéphane Dieterle, Nick Van Bakel, Kathrin Müller, Kirsten Sieverding, Jochen Weishaupt, Peter Munch Andersen, Markus Weber, Christoph Neuwirth, Markus Margelisch, Andreas Sommacal, Kristel R. van Eijk, Jan H Veldink, Project Mine ALS sequencing consortium, Géraldine Lautrette, Philippe Couratier, Tobias Boeckers, Albert C. Ludolph, Francesco Roselli, Deniz Yilmazer-Hanke, Stéphanie Millecamps, Edor Kabashi, Erik Storkebaum, Chantal Sellier, Luc Dupuis

**Affiliations:** Université de Strasbourg, Inserm, Mécanismes centraux et périphériques de la neurodégénérescence, UMR-S1118, Centre de Recherches en Biomédecine, Strasbourg, France; Department of Molecular Neurobiology, Donders Institute for Brain, Cognition and Behaviour and Faculty of Science, Radboud University, Nijmegen, Netherlands; Institute of Anatomy and Cell Biology, Ulm University, Ulm, Germany; German Center for Neurodegenerative Diseases (DZNE) Ulm, Ulm, Germany; Clinical Neuroanatomy, Department of Neurology, Ulm University, Ulm, Germany; Department of Neurology, Ulm University, Ulm, Germany; Laboratory of Translational Research for Neurological Disorders, Imagine Institute, Université de Paris, INSERM UMR 1163, 75015, Paris, France; Sorbonne Université, Institut du Cerveau - Paris Brain Institute - ICM, Inserm, CNRS, APHP, Hôpital de la Pitié Salpêtrière, Paris, France; Institute of Human Genetics, Ulm University, Ulm, Germany; Division for Neurodegenerative Diseases, Neurology Department, University Medicine Mannheim, Heidelberg University, Mannheim, Germany; Department of Clinical Science, Neurosciences, Umea; University, Umea, Sweden; Neuromuscular Disease Unit/ALS Clinic, Kantonsspital St. Gallen, St. Gallen, Switzerland; Institute for Pathology, Kanstonsspital St. Gallen, St. Gallen, Switzerland; Department of Neurology, University Medical Center Utrecht Brain Center, Utrecht University, Utrecht, The Netherlands; Service de Neurologie, Centre de Référence SLA et autres maladies du neurone moteur, CHU Dupuytren 1, Limoges, France

## Abstract

The genetic basis of amyotrophic lateral sclerosis (ALS) is still incompletely understood. Using two independent genetic strategies, we show here that a large part of ALS heritability lies in genes expressed in inhibitory and excitatory neurons, especially at splicing sites regulated by a defined set of RNA binding proteins including TDP-43 and FUS. We conducted a transcriptome wide association study (TWAS) and identified 59 loci associated with ALS, including 14 previously identified genes, some of them not previously reaching significance in genome wide association studies. Among the 45 novel genes, several genes are involved in pathways known to be affected in ALS such as mitochondrial metabolism (including *ATP5H, ATP5D, BCS1L*), proteostasis (including *COPS7A, G2E3, TMEM175, USP35*) or gene expression and RNA metabolism (including *ARID1B*, *ATXN3*, *PTBP2, TAF10*). Interestingly, decreased expression of *NUP50*, a constrained gene encoding a nuclear pore basket protein, was associated with ALS in TWAS (Zscore = −4, FDR = 0.034). 11 potentially pathogenic variants (CADD score > 20) in 23 patients were identified in the *NUP50* gene, most of them in the region of the protein mediating interaction with Importin alpha, and including 2 frameshift mutations. In cells from two patients carrying *NUP50* variants, we showed decreased levels of NUP50 protein. Importantly, knocking down *Nup50* led to increased neuronal death associated with p62 and nucleoporin inclusions in cultured neurons, and motor defects in *Drosophila* and zebrafish models. In all, our study identifies alterations in splicing in neurons as a critical pathogenic process in ALS, uncovers several new loci potentially contributing to ALS missing heritability, and provides genetic evidence linking nuclear pore defects to ALS.

## Introduction

Amyotrophic lateral sclerosis (ALS) is the most frequent adult onset motor neuron disease, and leads to death within a few years after onset through progressive paralysis caused by the simultaneous degeneration of upper motor neurons, located in the motor cortex, and of lower motor neurons in the brainstem and spinal cord. A variety of evidence has shown that ALS is a disease with high heritability. Twin studies^1^, prospective population based studies^2^ or register studies^3^ converged to an heritability ranging from 40 to 60%, while 5-10% of ALS patients show a family history^4, 5^. Genome wide association studies of ALS identified a hexanucleotide repeat expansion (HRE) in *C9ORF72* as a major ALS/FTD gene, and 15 genes are robustly associated to ALS in a cross ancestry manner^6, 7^, some of them having been found causative in familial ALS cases (eg *C9ORF72*, *SOD1, KIF5A* or *TBK1*). Despite these recent breakthroughs, a large part of heritability remains to be identified in ALS. Integration of proteome or transcriptome with GWAS studies, allowing identification of genes that confer risk to disease through protein or RNA abundance respectively, have yet to be performed in ALS to illuminate the nature of ALS heritability as recently exemplified in Alzheimer’s^8^ or Parkinson’s^9, 10^ diseases.

Beyond the identification of the gene variants associated to ALS, it would be critical to understand their functional relationship to well described pathogenic events. ALS is characterized by widespread occurrence of cytoplasmic aggregates of TDP-43, an RNA binding protein preventing aberrant inclusion of cryptic exons^11^. Interestingly, decreased TDP-43 function potentiates the inclusion of a cryptic exon in the *UNC13A* risk allele identified in GWAS, leading to decreased levels of the synaptic vesicle protein UNC13A, selectively in carriers of this allele^12, 13^. Whether other variants associated to ALS are also related to splicing events remains to be determined. Another critical pathogenic event in ALS is the dysfunction of the nuclear pore, what contributes to familial ALS caused by *C9ORF72* HRE^14–18^, lies also downstream of TDP-43 aggregation or loss of function^19–21^ and has been recently linked to *FUS*-ALS^22^. To date, there is however no genetic evidence directly linking core nuclear pore component(s) to ALS.

Here, we show that genes associated with ALS are enriched in neuronally expressed genes, and that ALS-associated variants are enriched in splicing regulating variants. Furthermore, a transcriptome wide association study identifies 45 new loci associated to ALS, most of them involved in pathways known to be pathogenically affected. In particular, loss of expression of *NUP50,* encoding a nuclear pore protein, was associated to ALS. Importantly, we also identified ALS patients carrying haploinsufficient mutations in *NUP50,* and demonstrate the critical role of NUP50 in neuronal survival and motor neuron function in multiple models.

## Results

### Cell type specific and molecular trait heritability of ALS

To better characterize ALS heritability, we applied stratified linkage disequilibrium ^23^ (LD) score regression (S-LDSC) to the ALS GWAS data^7^, to partition disease heritability using functional probabilistic annotations. To this aim, we computed the causal posterior probability (CPP) of each cis-SNP in the fine-mapped 95% credible set, using CAusal Variants Identification in Associated Regions (CAVIAR) fine-mapping method^24^. Then, for each SNP in the genome, we assigned an annotation value based on the maximum value of CPP across all phenotypes. We refer to this annotation as MaxCPP.

We compared ALS heritability estimates to two other neurodegenerative diseases (Alzheimer’s disease, AD, and Parkinson’s disease, PD) and two neuropsychiatric diseases (Autism spectrum disorder, ASD and schizophrenia, SCZ). Z-scores and p-values were calculated based on the coefficient *τ_c_* defined as the contribution of annotation *c* to per-SNP heritability for each annotation conditional on the baseline LD and the considered trait in order to test the unique contribution of each annotation.

We first asked whether heritability could be driven by genes expressed in specific CNS cell types. To answer this question, we used single-cell ATAC sequencing datasets of human-derived brain samples^25^, to specify cell-type specific gene expression based on open-chromatin region (OCR) and partitioned heritability into 6 different cell types. This analysis showed that ALS risk loci were significantly enriched for OCR in excitatory and inhibitory neurons (Bonferroni corrected p-value < 0.05), and to a lesser extent in oligodendrocytes (FDR < 0.05), but not in microglia or astrocytes (**Figure 1a, Supplemental Table 1**). The relevance of our analysis was corroborated by our observation of a significant enrichment of AD-risk loci in microglia-specific OCRs (**Figure 1a**) and of SCZ-risk loci in neuronal specific OCRs (**Figure 1a**) consistent with AD and SCZ underlying pathophysiology.

**Figure 1:**
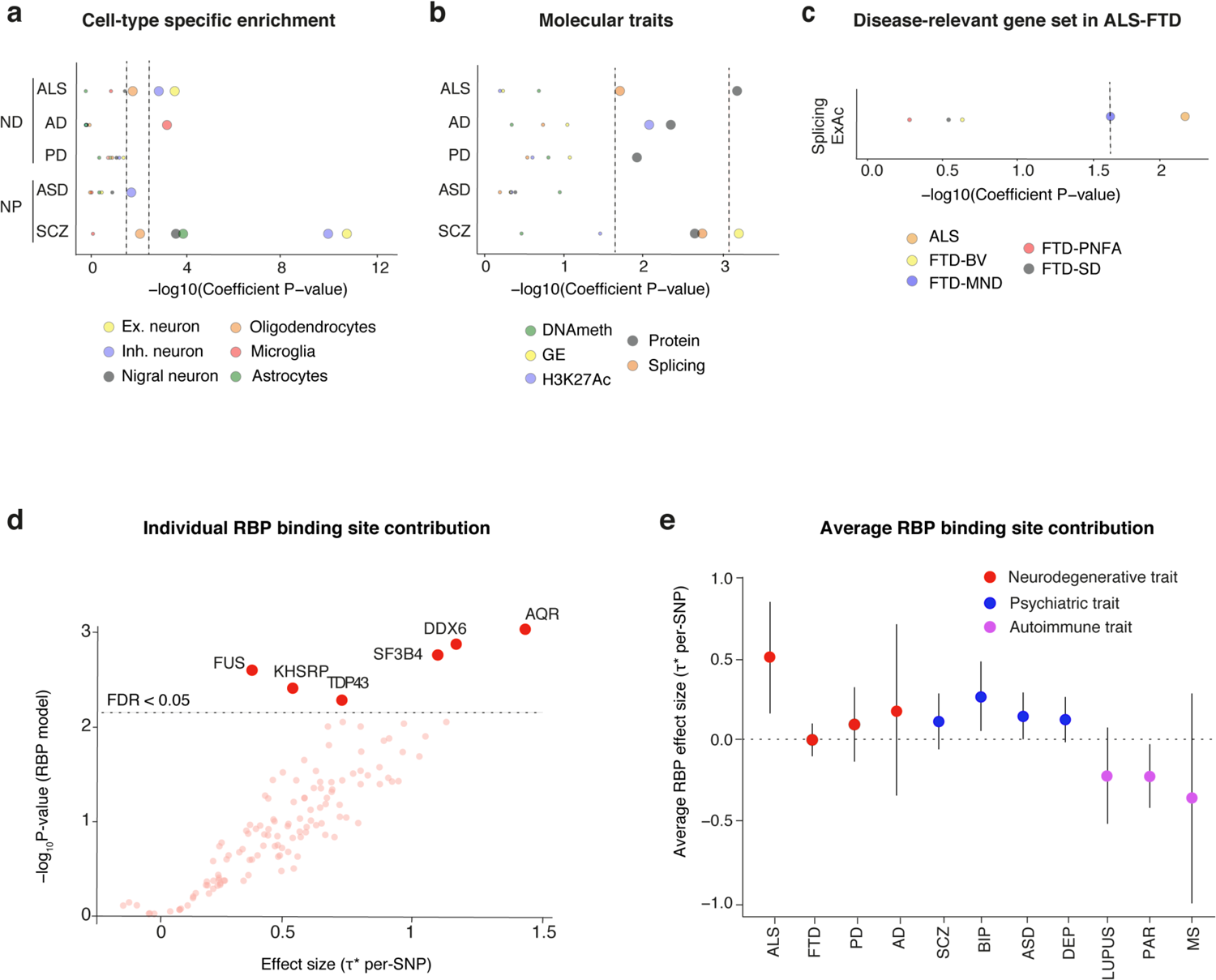
Cell type specific and molecular trait heritability of ALS. (a) Cell-type enrichment analyses indicate neuron-specific enrichment for inhibitory and excitatory neurons. No enrichment was found for microglia or other non-neuronal cell-types, contrasting the pattern observed in Alzheimer’s disease. Larger dots indicate statistically significant enrichment after Bonferroni correction p < 0.05. (b) LD score regression of molecular QTLs shows a significant enrichment in splicing and protein QTLs in ALS. We fit τ_c_ by conditioning on a collection of baseline annotations and all molecular QTLs to test the contribution of each individual annotation. Higher dashed lines indicates Bonferroni corrected p-values < 0.05 while lower dashed lines indicates FDR < 0.05. (c) Disease-relevant genes sets analysis shows a significant enrichment of splicing QTLs in genes depleted of protein truncating variant in ALS and FTD-MND but not in other FTD subtypes. Larger dots indicate statistically significant enrichment after Benjamini-Hochberg correction FDR <= 0.05. (d) The per-SNP heritability effect sizes (τ*) for each RBP target site dysregulation is plotted for ALS GWAS. The dashed line indicates RBP models FDR < 0.05 threshold after multiple hypothesis correction (block jackknife-based one-sided p-values; Benjamini-Hochberg correction). (e) The per-SNP heritability effect sizes (τ*) for RBPs after conditioning on a collection of molecular QTL annotations (i.e. independent RBP effects from molecular QTLs and baseline annotations). All error bars are 95% CI. ALS = amyotrophic lateral sclerosis, FTD = Fronto-Temporal Dementia, PD = Parkinson’s disease, AD = Alzheimer’s disease, SCZ= Schizophrenia, DEP = Depression, BIP = Bipolar Disorder, ASD = Autism Spectrum Disorder, PAR = Rheumatoid Arthritis, MS = multiple sclerosis

We then asked whether fine-mapped posterior probabilities for several different molecular QTLs are strongly enriched for ALS heritability. To this aim, we constructed MaxCPP annotations using brain specific eQTL, meQTL, sQTL and pQTL and used previously described hQTL from the BLUEPRINT consortium^26^. We included a broad set of 75 functional annotations from the baseline LD model in most analyses. We partitioned disease heritability on 5 molecular traits and assessed statistical significance at FDR < 0.05 after Benjamini-Hochberg correction. Here again this analysis yielded expected results such as the observed significant enrichment for enhancer (H3K27Ac) and protein QTLs in Alzheimer’s disease (**Figure 1b, Supplemental Table 2**) which is supported by a prominent role of epigenetic modification and protein expression in this disorder^27^. Most importantly, splicing QTLs were enriched in ALS heritability as well as in SCZ, but not in other diseases (AD, PD, ASD) (**Figure 1b, Supplemental Table 2**), consistent with a selective role of splicing dysregulation in ALS and SCZ. To strengthen this potential role of splicing dysregulation in ALS heritability, we used the ExAC gene set, which consists of 3,230 genes that are strongly depleted of protein-truncating variants^28^, and constructed a new splicing QTLs annotation named (MaxCPP-ExAC), defined as the maximum CPP restricted to genes in the ExAC gene set. Conditional analysis on the baseline LD model and all QTLs showed a significant enrichment for splicing QTLs in genes depleted of protein truncated variants in ALS as well as FTD-MND but not in forms of FTD devoid of motor neuron involvement (**Figure 1c, Supplemental Table 3).** suggesting that the enrichment of risk loci in splicing QTLs annotation is specific to motor neuron disease.

Having established the importance of splicing regulation in ALS, we next investigated the potential role of individual RNA-binding protein (RBP) dysregulation^29^. To address this hypothesis, we applied the statistical framework of stratified LD score regression^79^ to partition disease heritability into the annotated binding sites of 125 RBPs. The LD score regression framework allows estimation of SNP effects (τ*) standardized for comparison across different disease or trait GWAS studies while conditioning on a baseline functional annotation. Importantly, we observed significantly elevated effect size (τ*) estimates in 6 RBPs after correcting for multiple hypothesis testing (**Figure 1d**, FDR < 0.05, Benjamini-Hochberg correction, **Supplemental table 4**). Strikingly, among these 6 RBPs, we observed disrupted target sites of TDP-43 (τ* =0.72; p-jackknife = 2.9×10-3) and FUS (τ* = 0.36, p = 2.6×10-3, jackknife), that contributed significantly to ALS heritability, but also associations with novel RBPs such as KHSRP (τ* = 0.52, p = 4.2×10-3, jackknife) or AQR (τ * = 1.44, p = 9.9×10-4, jackknife). To determine the selectivity of this involvement of RBP dysregulation in ALS, we compared global RBP binding sites dysregulation in 11 different disease traits. To this aim, we first estimated the effect size of individual RBP for each disorder while jointly conditioning on the MaxCPP QTL-based annotations and then averaged τ* across the 125 RBPs tested in our models. We found that ALS displays the highest average RBP effect size τ* across the 11 traits (averaged τ* = 0.506, sd = 0.1729, **Figure 1e**, **Supplemental table 5**) suggesting a prominent contribution of disrupted RBP binding sites to ALS heritability. Overall these data suggest that ALS heritability is significantly caused by perturbations of splicing regulation in particular at TDP-43 and FUS binding sites.

### Polygenic risk score analysis suggests that splicing defects in inhibitory neurons confer ALS risk

We sought to independently confirm our results by performing polygenic risk score analysis^30^. For this, we used 3 independent datasets, the reference dataset (12,577 ALS cases and 23,475 controls) (reference), the training dataset (5226 ALS cases and 7226 controls) and the replication dataset (2298 ALS cases and 3414 controls) (**Figure 2a).** The training and the replication sets are individual-level genotype and phenotype data (accession number phs000101.v5.p1). The molecular QTLs that achieved significance in the training dataset (false discovery rate (FDR)-corrected p-value < 0.05) were selected for replication. Then, we tested whether this set of QTLs achieved significance in the replication dataset (raw p-value < 0.05). In order to maximize sensitivity in our analysis, we restricted our test SNPs to the SNPs with a MaxCPP value > 0.9 in each annotation, which have a high posterior probability of being causal for any given molecular QTLs and then calculated the contribution to ALS risk of 5 major molecular QTLs (**Supplemental table 6**). Importantly, this strategy identified that splicing QTLs were significantly associated with ALS in the training dataset (**Figure 2b**) (FDR = *0.032). This was independently confirmed in the replication dataset, with ALS risk being significantly associated with sQTLs in ALS patients non carrying a *C9ORF72* HRE (**Figure 2c).** Then, we wanted to test whether a significant risk associated with ALS could be associated to cell-type specific splicing defects. To do so, we first extracted OCR from 12 independent cell-types and tested each annotation corresponding to a specific cell-type for its contribution to ALS risk. We observed that 5 significant cell-types, all neuronal, were significantly associated with ALS (**Supplemental table 7).** We then intersected SNPs for each cell type annotation to the sQTLs and tested this new set of SNPs in our replication dataset. We observed that sQTLs in somatostatin interneurons were significantly associated with ALS risk (**Figure 2d).** Overall, our results show that variants associated with splicing regulation in inhibitory neurons contribute to ALS risk.

**Figure 2:**
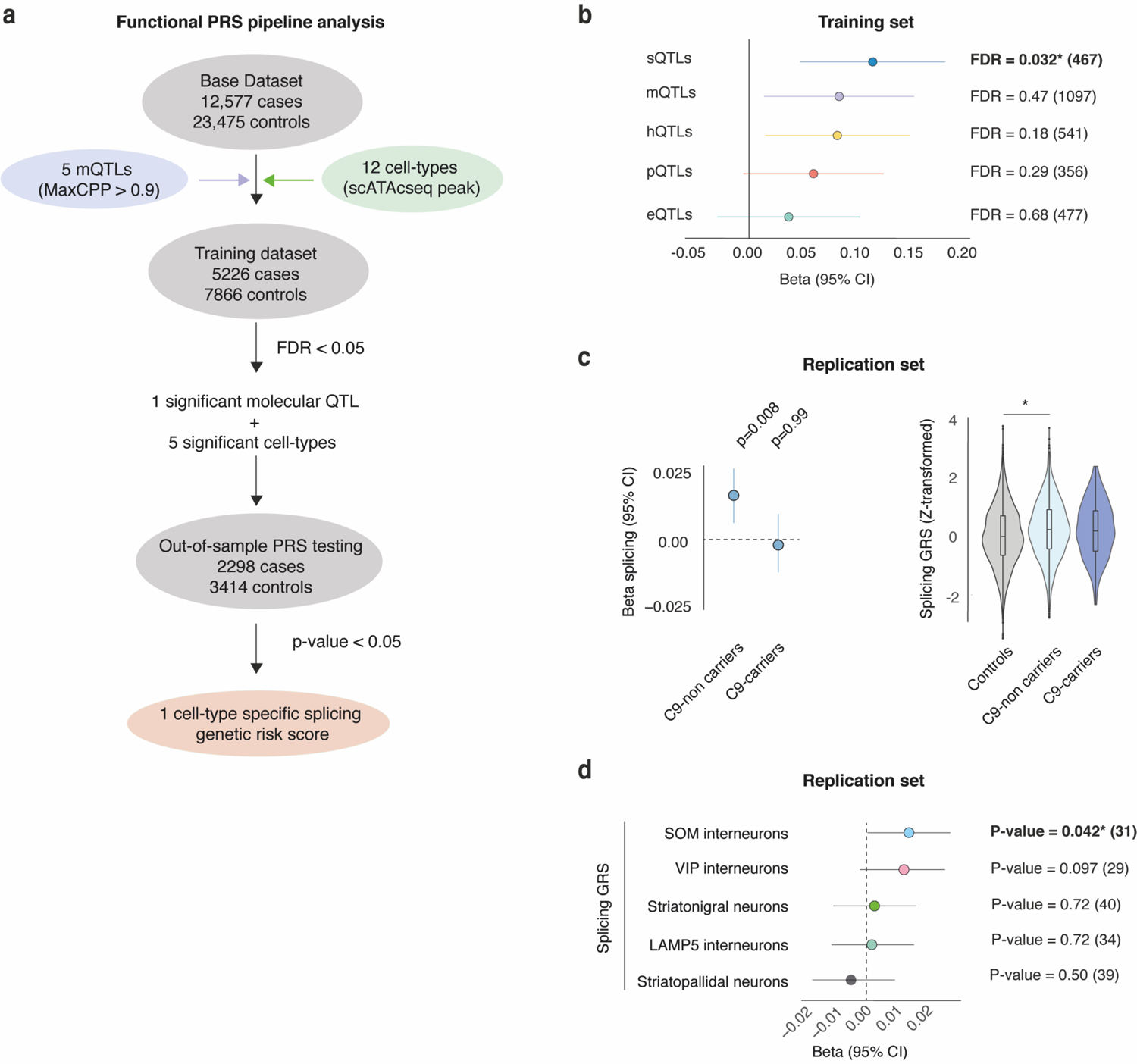
Polygenic risk score identifies molecular QTLs and cell types contributing to the risk of developing ALS. Analytic pipeline. (**a**) The Forrest plots show polygenic risk score estimates in the training set for each molecular traits. * asterisk denotes molecular QTLs that reached genome-wide significance (FDR < 0.05) and variants left after the clumping procedure in each category are shown in between brackets. (**b**) Logistic regressions were performed to evaluate the association between the molecular QTLs polygenic risk score of interest with ALS as the outcome, glm (PHENO ∼ zSCORE) in C9 carriers and C9 non-carriers. Beta coefficients were used to derive p-value for each condition. * denotes genome-wide adjusted significance p < 1.10e-5 (**c**) Variants in significantly associated cell-type were intersected with splicing QTLs and tested in the replication set. * denotes genome-wide significance p < 0.05. The Forest plot displays the distribution of beta estimates across molecular QTLs, with the horizontal lines corresponding to 95% CI.

### TWAS of ALS identifies *NUP50* loss of expression as a risk factor for ALS

To identify and prioritize candidate ALS genes, we performed TWAS^31^ using summary-level data from the largest ALS GWAS of European ancestry (29,621 cases and 120,971 controls)^7^ and transcriptome panels from dorso-lateral prefrontal cortex (DLPFC). The TWAS approach uses information from the RNA expression and splicing measured in a reference panel and the ALS GWAS summary statistics to evaluate the association between the genetic component of expression and ALS status (see Methods). We built TWAS models using transcript expression data from DLPFC RNA-seq data (*n* = 551) from BrainSeq phase 2. In addition to using transcript expression to build predictive models, RNA-seq data from DLFPC allowed us to consider alternative splicing in ALS by quantifying “percent spliced in” of splicing events^32^ in the CMC dataset (n=431) and the Mayo dataset (n=231). We used the FUSION software to estimate the heritability, build predictive models, and perform TWAS. For each reference panel, FUSION estimates the heritability of transcript expression and alternative splicing explained by local SNPs (i.e., 1 Mb from TSS of each gene) using linear-mixed models.

Transcripts or splicing events that are nominally significant at *p*< 0.01 for SNP heritability (*cis-h_g_*^2^) are used for training predictive models. FUSION fits four predictive linear models (see Methods) for every gene or intronic excision event using local SNPs as predictors. The models with the best cross-validation prediction accuracy are kept for prediction into GWAS. In total, 18,653 transcripts, and 20,370 DLPFC junctions, were used for TWAS in the BrainSeq dataset. For the CMC and the Mayo dataset, 7,695 and 12,280 spliced introns were used for TWAS association, respectively. The square of correlation (*R*^2^) between predicted and observed gene expression levels normalized by corresponding *cis*-*h_g_*^2^ was calculated to measure prediction accuracy. Across all cohorts least absolute shrinkage and selection operator (LASSO) attained the best predictive performance, with 30% improvement in prediction *R*^2^ over other models (**Supplemental figure 1**). Using these TWAS models, we found that 59 genes are significantly associated with ALS in DLPFC or hippocampus through RNA expression and/or splicing (FDR < 0.05) (**Figure 3a**, **Supplemental table 8, Supplemental table 9**). 14 out of the 59 unique TWAS genes are in known ALS GWAS susceptibility loci, including *C9ORF72, MOBP, NEK1* or *SCFD1* leaving 45 potential novel associations. Among the novel associations we observed genes previously associated with Parkinson’s disease such as *TMEM175* (conditional *p* < 2.20 × 10^−6^, **Supplemental table 10 and Supplemental figure 2**) and *IDUA* (conditional p.< 3.20 × 10^−5^, **Supplemental table 10**) suggesting common underlying mechanisms in both disorders.

**Figure 3.**
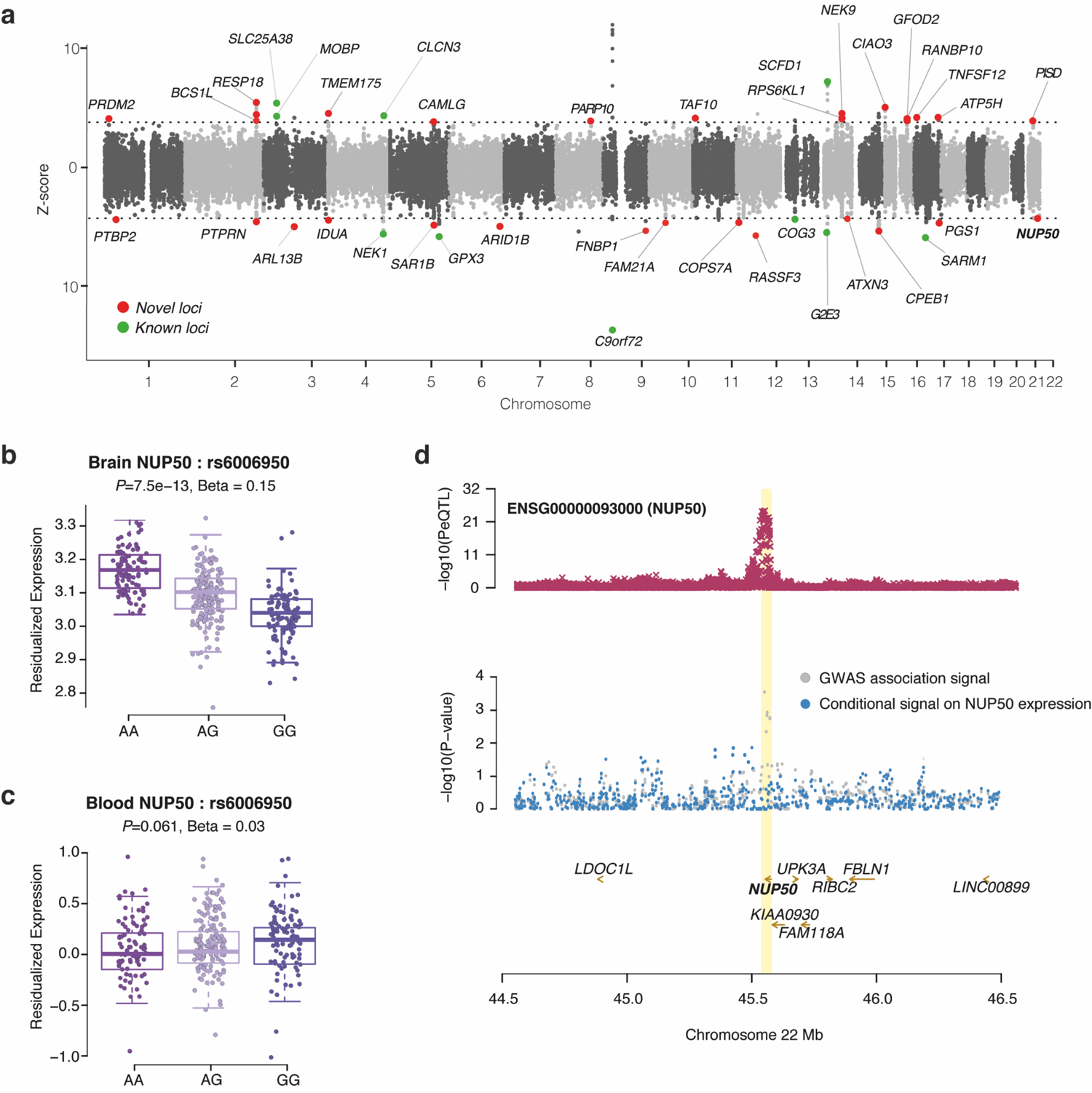
Transcriptome-wide association study of Amyotrophic Lateral Sclerosis. (**a**) Manhattan plot of ALS transcriptome-wide association study (TWAS) using gene expression and splicing models from dorso-lateral prefrontal cortex (DLPFC). Each point represents a single gene tested, with physical position plotted on the *x*-axis and *Z*-score of association between gene expression or intronic splicing with PD plotted on the *y*-axis. Colored (black or blue) points represent significant association to ALS at a suggesting FDR < 0.05. (**b-c**) Boxplot showing the association between a single-nucleotide polymorphism (SNP) (rs6006950) that tags the ALS genome-wide association studies (GWAS) risk loci at *NUP50* and gene expression level of *NUP50* in DLPFC (b) and blood from the GTEx consortium (c). rs6006950 is significantly associated with the expression level of *NUP50* in DLPFC, but not in blood. (**d**) LocusZoom style plot for the region surrounding *NUP50* shows colocalization of the DLPFC *NUP50* expression quantitative loci (eQTL) (middle panel) and ALS GWAS association signal (top). ALS TWAS signal at the *NUP50* locus (gray) and TWAS signal after removing the effect of *NUP50* expression (cyan). This analysis shows that the association is largely explained by *NUP50* expression.

Most of the novel associated genes fell into functional categories previously associated with ALS^4,5^. These categories included mitochondrial energy metabolism, with two genes encoding subunits of the mitochondrial ATP synthase (*ATP5H,* conditional *p* < 1.10 × 10^−5^ and *ATP5PD* conditional *p* <7.6 × 10^−7^) and *BCSL1* conditional *p* < 3.10 × 10^−5^, **Supplemental table 10**), encoding a chaperone for mitochondrial cytochrome c reductase. Consistent with a genetic involvement of proteostasis defects in ALS, we also observed novel associations with *COPS7A* (conditional *p* < 1.4× 10^−5^) and *USP35* (conditional *p* < 1.2 × 10^−6^, **Supplemental table 10**) two genes involved in ubiquitin mediated proteolysis.

A major result of our TWAS was that as many as 14 novel associated genes were, directly or indirectly, associated with gene expression and RNA metabolism. Illustrating this, we found one novel association with ALS in *CPEB1* (conditional *p* < 7.70 × 10^−7^, **Supplemental figure 3**), *RANBP10* (conditional *p* < 1.70 × 10^−5^, **Supplemental table 10**) and *TAF10* (conditional *p* < 1.40 × 10^−5^, **Supplemental figure 4, Supplemental table 10**). Most interestingly, we also found a significant association with the RNA-binding protein *PTBP2* in the hippocampus dataset (conditional *p* < 5.80 × 10^−5^, **Supplemental figure 5, Supplemental table 10**). This candidate appears especially relevant since PTBP2 is an RNA binding protein that has been shown to mediate axonogenesis-associated splicing during neurogenesis and is essential for the repression of cryptic exons in differentiated neurons ^33, 34^. Interestingly, a large proportion of the genes identified as involved in gene expression or splicing also displayed a pLI > 0.9 suggesting that they are highly intolerant to the loss of function^28^ reinforcing the idea that ALS-causing variants could be enriched in highly-intolerant genes associated with transcriptional regulation and splicing ^35^. Highly significant association (FDR < 0.05) with gene expression were replicated in independent datasets and tissues (**Supplemental figure 6**), further confirming the relevance of this class of genes. Finally, we summarized findings from 6 independent TWAS studies recently published^9, 36–38, 39, 40^. For each individual dataset, significant TWAS genes were used to compute the average pLI (probability of loss of function intolerance) score. We observed a significant increase in the average pLI score for the TWAS ALS genes compared other neurodegenerative disorders such as Parkinson’s disease (pLI ∼ trait, F_(6,301)_ = 2.665; *p = 0.0167) or bvFTD (pLI ∼ trait, F_(6,301)_ = 2.665; *p = 0.0303) (**Supplemental figure 7a**). Also, we found that almost ∼37% of the TWAS ALS genes have a pLI score > 0.9 suggesting a prominent role of highly constrained genes in ALS pathogenesis (**Supplemental figure 7b**).

**Figure 4:**
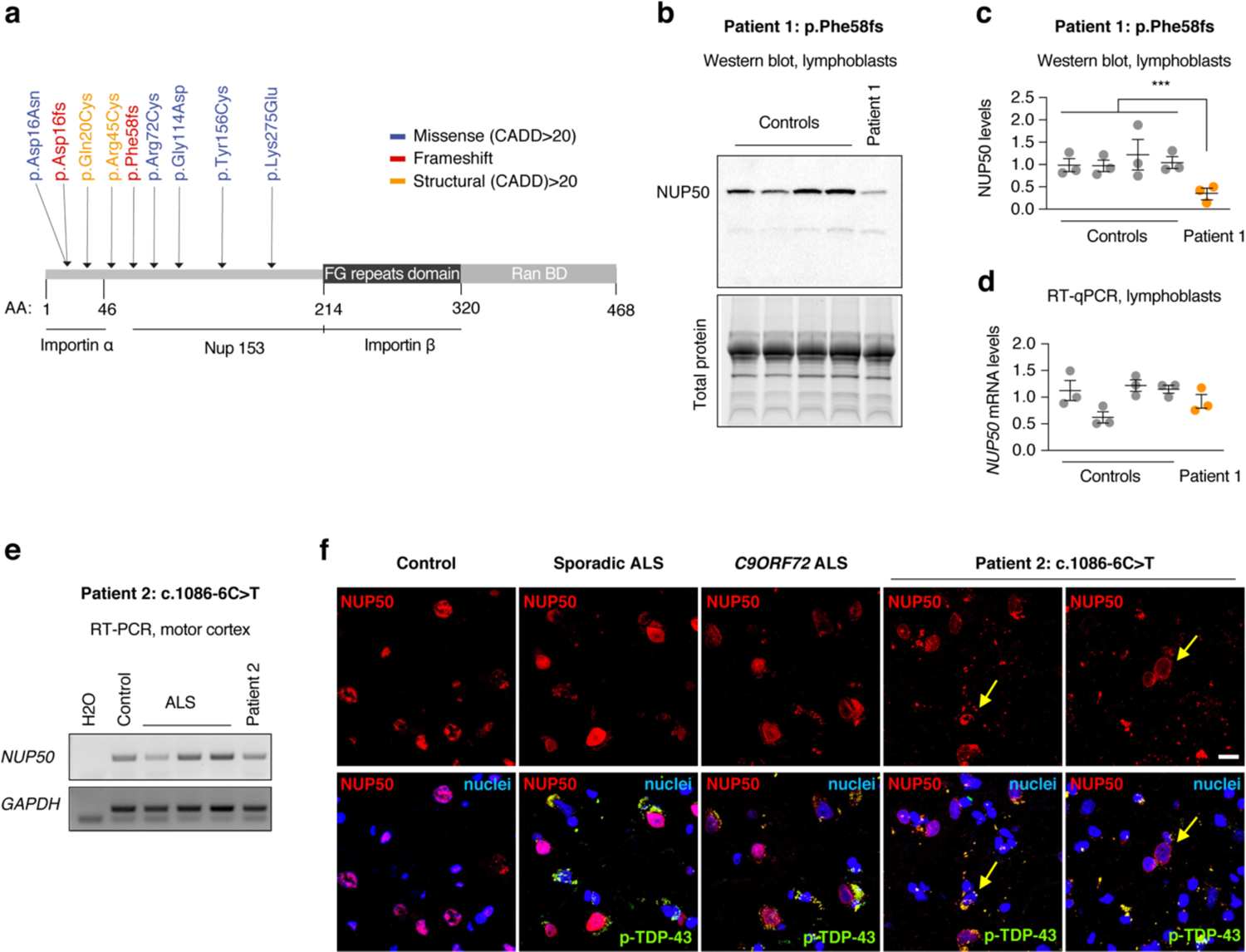
*NUP50* variants in ALS. (**a**) High confidence NUP50 variants with a CADD (combined annotation-dependent depletion) score >20 are indicated. Most of these variants are located in or near the importin alpha domain of the NUP50 protein suggesting a role in nucleo-cytoplasmic transport. (**b-d**) NUP50 expression analysis in lymphoblasts from healthy donors and patient 1 carrying the NUP50 frameshift Phe58fs mutation. Western blotting (b,c) shows a significant decrease in NUP50 protein levels (One way ANOVA: F_(1,13)_ = 12.5, *p* = 0.00365, Control ∼ Patient: post-hoc Tukey, adjusted-*p*= **0.00367). However, we observed no significant changes in *NUP50* mRNA expression (d, Two-sample t-test t = 0.20955, *p* = 0.8348). (**e**) RT-PCR of motor cortex extracts from non-neurological control, 3 ALS cases and patient 2 carrying the near-splice NUP50 mutation c.1086-6C>T shows decreased *NUP50* RNA levels. GAPDH is used as a loading control. (**f**) Immunofluorescence analysis of NUP53 and phospho-TDP-43 in the same cases as in (e). Please note the NUP50 pathology in Patient 2 (arrows).

**Figure 5:**
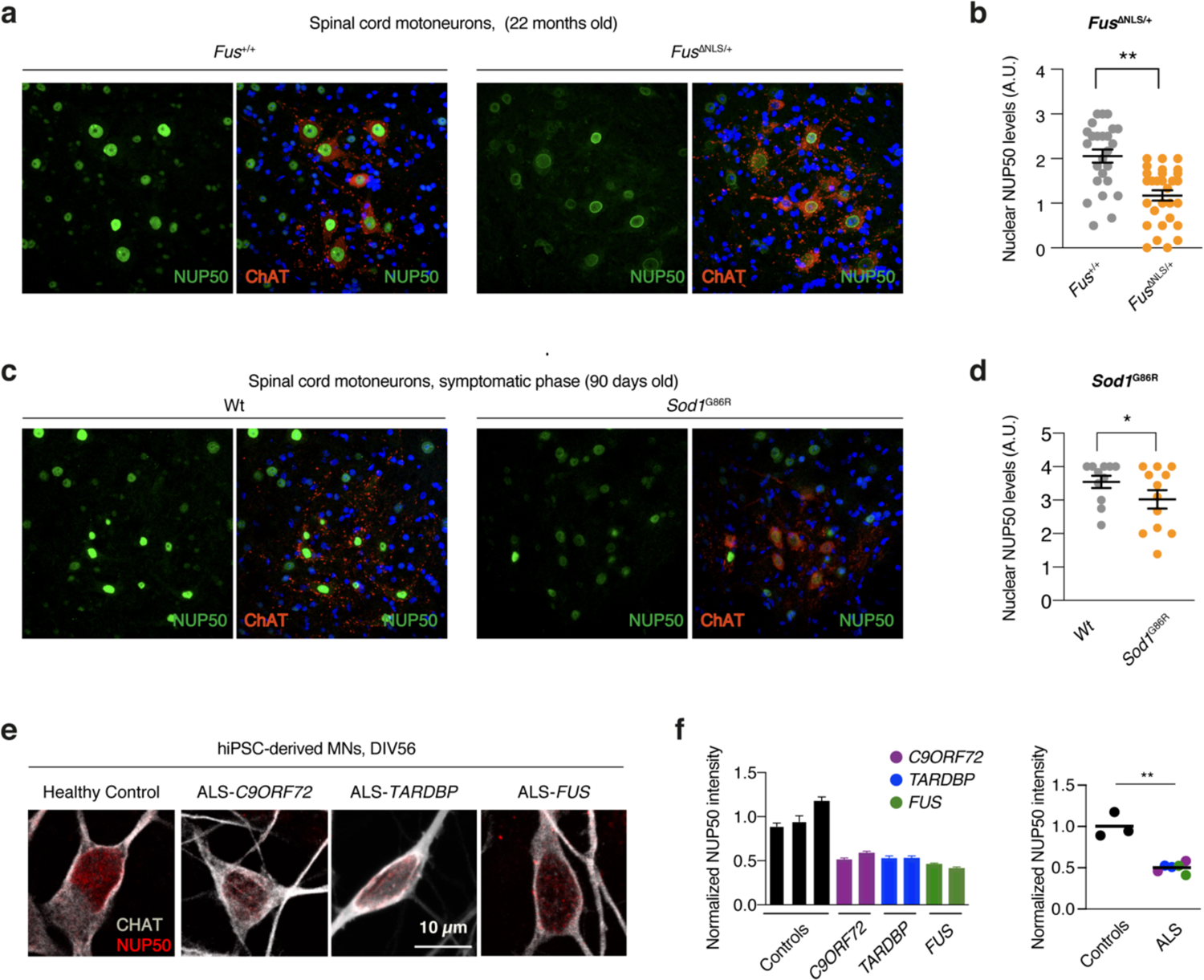
NUP50 nuclear loss is a unifying feature in cell and animal models of ALS. (a-d) Representative image of immunohistochemistry for NUP50 and CHAT in spinal cord sections showing a decrease in nuclear NUP50 levels in motor neurons of (a-b) Fus^ΔNLS/+^ (Nested-t-test: t=3,293, df=10, *p* = **0,0081) and (**c-d**) Sod^G86R^ (Nested-t-test: t=2,212, df=19, *p* = *0.038) mice. (**e**) Representative image of iPSCs-derived motoneurons from fALS (FUS, TDP-43 and C9ORF72 HRE carriers) immunostained for CHAT (white) and NUP50 (red). (**f**) Quantification of NUP50 levels shows a significant decrease in all fALS mutation carriers (Unpaired t-test: t=5,552, df=2,320, *p* = *0.0220).

**Figure 6:**
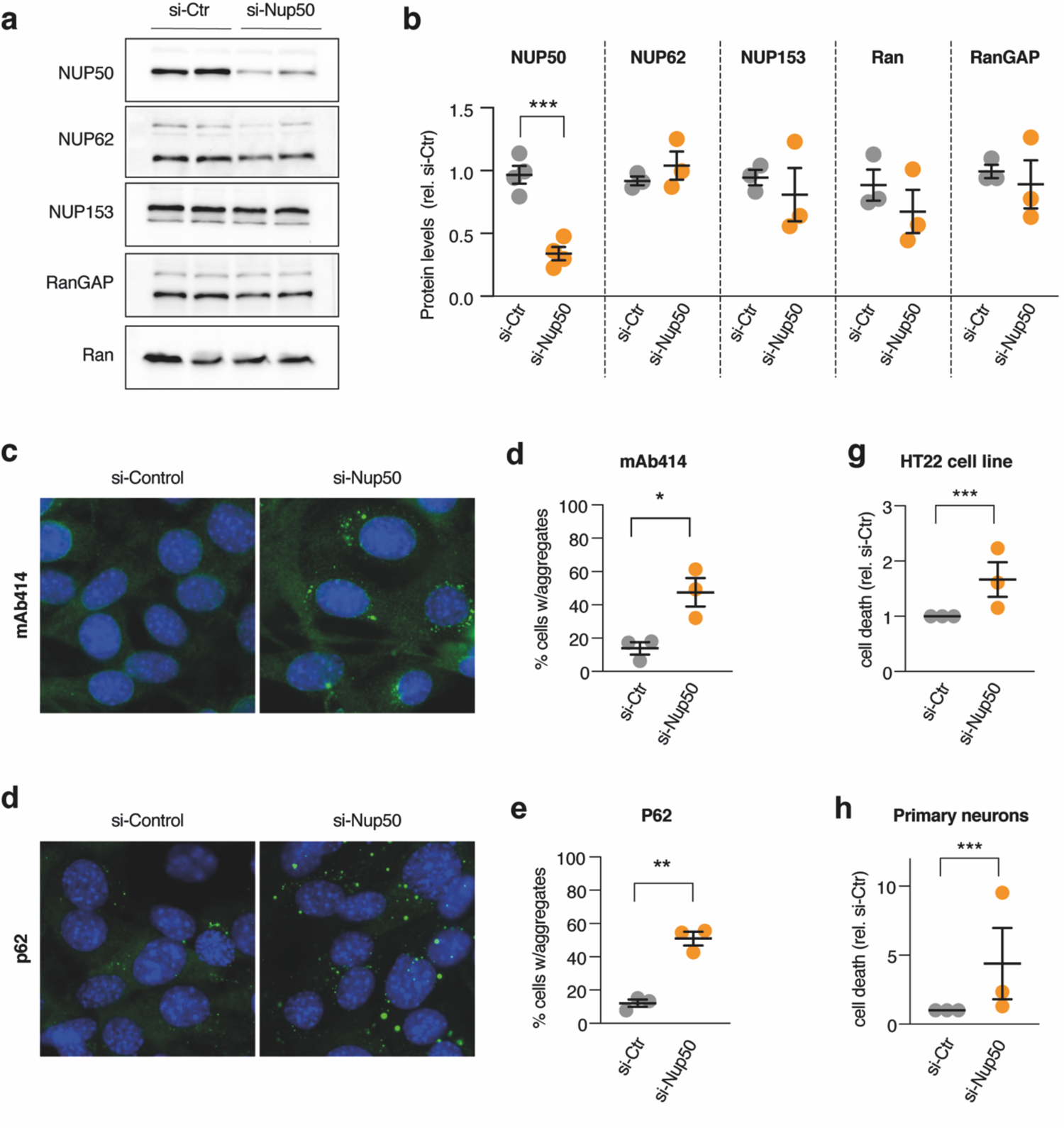
knockdown of NUP50 in mouse neuronal cells leads to cell death and ALS-like pathological features. (**a**) Representative images of western blots for NUP50 and different nucleoporins (**b**) Dot-plots showing a decrease in NUP50 levels (Nested-t-test: t=8,124, df=14, \*\*\*\**p* < 0.0001) but not other associated nucleoporins and RanGAP (p > 0.05) after knock-down of the *Nup50* mRNA (si-Nup50) compared to the control condition. (**c) (d**) Representative images and dot plots showing an increase in cytoplasmic inclusions of nucleoporins as stained with mAb414 recognizing the repeated FXFG repeat sequence in nucleoporins in HT22 cell line upon Nup50 knock-down (Nested-t-test: t=6,778, df=16,****p<0.0001). (**e) (f**) Representative images and dot plots showing an increase in p62 inclusion in HT22 cell line upon Nup50 knock-down (Nested-t-test: t=9,846, df=17,****p<0.0001). Dot-plots showing a significant increase in neuronal death (**g**) in HT22 cell lines (Nested-t-test: t=3,721, df=24, **p=0.0011) and in mouse primary neurons (**h**) (Nested-t-test: t=3,18, df=34, **p=0.0031)

**Figure 7:**
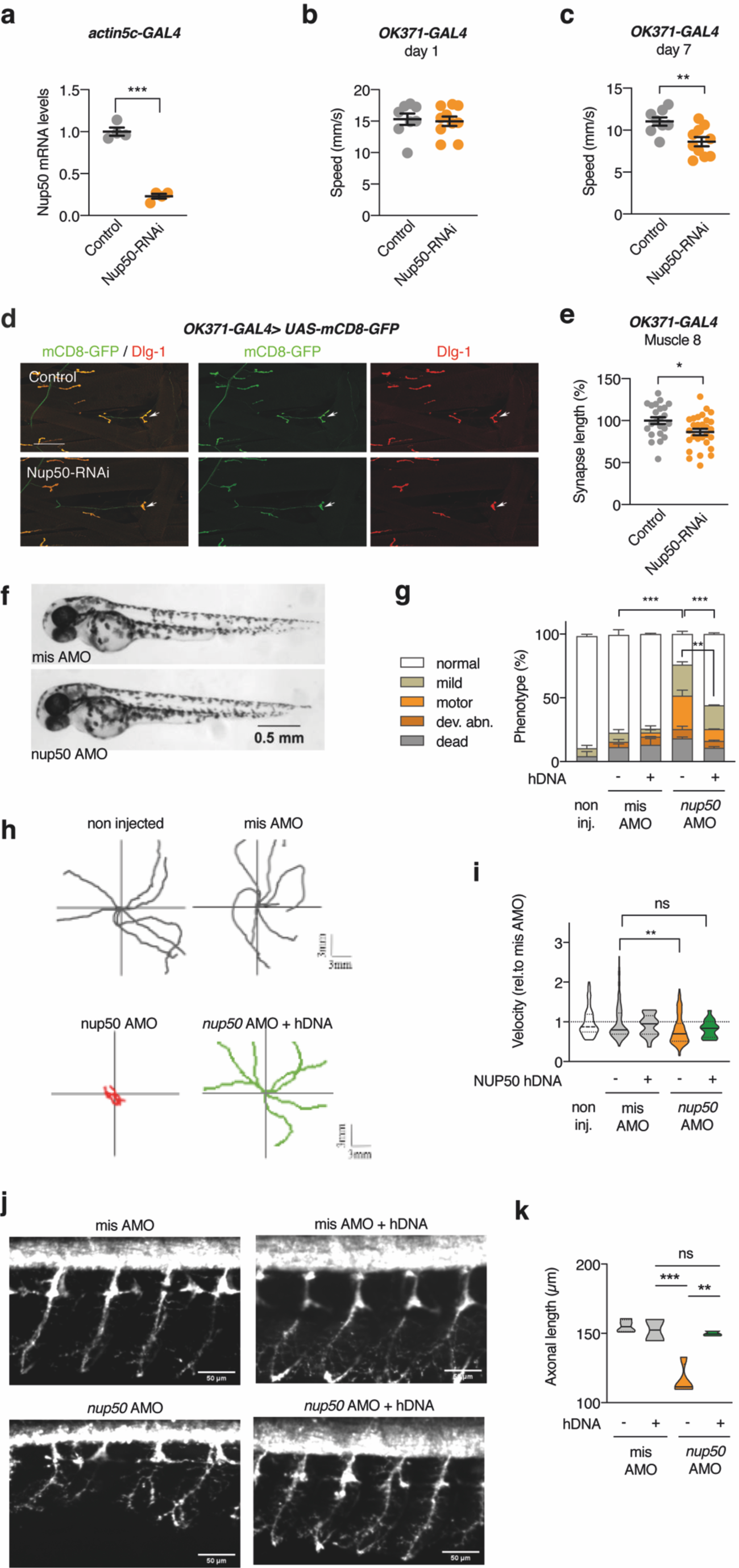
knockdown of Nup50 leads to motor defects in Drosophila and Zebrafish. (a-e) Effect of *Drosophila Nup50* knock-down on motor performance and NMJ morphology. (**a),** mRNA expression level (% of driver-only control) of Nup50 in third instar larvae ubiquitously (*actin5C-GAL4*) expressing Nup50-RNAi. n=4 per genotype; ***p<0.0001 by unpaired t-test. (**b),(c),** Climbing speed in an automated negative geotaxis assay of 1-(b) or 7-day-old (c) male flies expressing Nup50-RNAi selectively in motor neurons (*OK371-GAL4*). n=8-10 groups of 10 flies per genotype; **p<0.01 by unpaired t-test. (**d),** Representative images of the NMJ on muscle 8 (white arrow) as visualized by the expression of membrane GFP (*UAS-mCD8-GFP*) in motor neurons (*OK371-GAL4*) which shows axons and the presynaptic compartment of NMJs (green), and dlg1 immunostaining labelling the postsynaptic compartment of NMJs (red) in third instar larvae expressing Nup50-RNAi selectively in motor neurons (*OK371-GAL4*) compared to the driver-only control. Scale bar: 100 μm. (**e),** Synapse length on distal muscle 8 of third instar larvae expressing Nup50-RNAi selectively in motor neurons (*OK371-GAL4*). n=23-28 per genotype; *p<0.05 by unpaired t-test. (**f**) Representative images of 48 hpf zebrafish embryos showing no difference on global morphology. (**g**) Distribution of motor phenotypes (average percent of the group) of 48 hpf zebrafish embryos among the different conditions. Data are presented as mean +sem; **p<0,01 and ****p<0,0001 by Tukey’s multiple comparison test following 2way ANOVA. 1. (**g**) Representative swimming trajectories of 48 hpf zebrafish embryos upon TEER test. 2. (**h**) Quantification of average swimming velocity of 48 hpf zebrafish embryos. Each dot represents one embryo. Data are normalized to the respective mis AMO control and presented as mean +sem; **p<0,01 and non-significant (*ns*) by Tukey’s multiple comparison test following 1way ANOVA. (**i**) Representative images of spinal motor neurons of 48 hpf zebrafish embryos. 3. (**j**) Quantification of average motor neuron axonal lengths of 48 hpf embryos. Data are presented as mean +sem; **p<0,01, ***p<0,001 and non-significant (*ns*) by Tukey’s multiple comparison test following 1way ANOVA.

Among the genes associated to gene expression and splicing, we observed a novel association between ALS and the decreased expression of *NUP50* (Zscore = −4, FDR = 0.034*, **Figure 3a**), encoding a nuclear pore basket protein. Also, we found that the ALS risk allele (rs6006950-G) at the *NUP50* locus is associated with decreased expression of *NUP50* in the DLPFC (rs6006950, *p* = 7.5 × 10^−13^), but not in the blood (*p* = 0.061) (**Figure 3b).** Using FUSION we found that only one expression event (decreased expression of NUP50) remained significant after conditioning on all other association signals (conditional *p* < 6.60 × 10^−5^, FDR = 0.034; **Figure 3d**, **Supplemental table 10**). These results suggest that loss of expression of *NUP50* in the central nervous system could constitute a risk factor for ALS. Since there was, to our knowledge, no previous genetic evidence linking the core components of the nuclear pore to ALS, we selected *NUP50* for further genetic and functional analysis.

### Identification of rare frameshift mutations in *NUP50* causing haploinsufficiency in ALS

Since *NUP50* is a constrained gene according to the LOEUF metrics quantified by the gnomAD group^41, 42^, we investigated how rare, coding variants of NUP50 contribute to ALS risk in whole-genome sequencing dataset of ALS patients (N = 6,596) and controls (N = 2,454) through exome-wide association analysis, including transcript-level rare-variant burden testing for different models of allele-frequency thresholds and variant annotations^7^. Most *NUP50* derived transcripts, including the canonical *NUP50* transcript (ENST00000347635), displayed a suggestive p-value of 0.025 for damaging variants at MAF < 0.01 and MAF < 0.005 and a p-value of 0.050 for disrupting and damaging variants at MAF < 0.01 and MAF < 0.005. To characterize the potential *NUP50* variants, we filtered all single-nucleotide polymorphisms (SNP) and short coding insertions/deletions affecting coding sequence, splice acceptor/donor site or non-coding regions (introns, 5’UTR, 3’UTR) of *NUP50*, and identified 11 potentially pathogenic *NUP50* variants in Project Mine database as well as in French^43^ and German^44^ exome datasets in 23 patients (**Figure 4a, Table 1**). Among the 9 missense and structural mutations identified across the different cohorts, 7 have CADD (combined annotation dependent depletion) scores > 20 (**Table 1**) suggesting high pathogenicity. However some of the missense *NUP50* mutations identified in ALS patients were also reported at very low frequency in the ExAC database (minor allele frequency<0.0003) classifying them rather as strong ALS risk factors.

Among these variants, we focused our attention on two frameshift mutations, that possibly lead to haploinsufficiency. Lymphoblasts were available for Patient 1, carrying a single nucleotide deletion (c.174del) leading to frameshift and premature stop codon (p.Phe58Leufs*37), which was confirmed by Sanger sequencing (**Supplementalfigure 8a-b**). Other ALS genes were further interrogated for this specific patient, and did not show potentially pathogenic variants, as variants in *SS18L1* (c.1036+17_1036+133del) and *NEFH* (c.85G>T, p.Ala29Ser) were present in control databases. Patient 1 had disease onset in his 30’s with signs of upper and lower motor neuron involvement first restricted to the lower limbs, leading to a slowly progressive paralysis. He died from pulmonary embolism after bacterial meningitis in his 50’s. Western blotting showed decreased levels of NUP50 protein in lymphoblasts of Patient 1 as compared to 4 healthy controls (**Figure 4b-c**), while *NUP50* mRNA levels were unchanged (**Figure 4b**), consistent with the detection of the mutant allele expression after cDNA sequencing (**Supplementalfigure 8a-b**).

An autopsy was available from the fALS patient carrying the novel *NUP50* nearsplice mutation c.1086-6C>T immediately upstream of exon 7 (Patient 2). Exome-sequencing and *C9ORF72* repeat-primed PCR did not reveal any rare mutations in other genes associated with ALS. To evaluate the consequences of this substitution, RT-PCR of *post mortem* motor cortex samples was performed using oligonucleotides spanning exons 5 to 8 of NUP50 mRNA. While no aberrantly spliced products could be detected, semi-quantitative RT-PCR indicated decreased expression of *NUP50* mRNA (**Figure 4e**). Since the allele carrying the *NUP50* nearsplice mutation c.1086-6C>T also carried an inframe deletion (rs773541780, c.1065_1067delAGA; p.Glu355del, MAF in ALS patients: 1.15E-04; MAF in non-finnish European population: 1.465E-04, **Supplementary Figure 9a-b**), we were able to discriminate expression of both alleles and cDNA sequencing confirmed lack of expression of the *NUP50* nearsplice mutation c.1086-6C>T allele (**Supplementary Figure 9c**). Intriguingly, a similar downregulation of NUP50 was observed in one of the 3 ALS patients included as control (ALS 1 in **Figure 4e**). This patient did not carry mutations in most known ALS genes or rare variants of interest in *NUP50* exons and splice sites. Immunofluorescence analysis showed decreased levels of NUP50 in neurons and widespread ring-shaped perinuclear neuronal NUP50 labeling in Patient 2 (arrows in **Figure 4f**), while NUP50 pathology was also observed in TDP-43 positive cells in one sporadic and one *C9ORF72* HRE patient analyzed in parallel (**Figure 4f**). Semi-quantitative analysis of NUP50 immunoreactivity further confirmed that NUP50 immunoreactivity was generally lost in ALS samples^45^ as compared to a non neurological control, but this loss was more pronounced in Patient 2 (**Supplementary Figure 10**). Together, these results show that rare *NUP50* mutations are occurring in ALS patients, leading to loss of NUP50 protein.

### Loss of nuclear NUP50 is a unifying feature in ALS patients-derived cell lines and animal models

We then sought to determine the effect of familial ALS mutations on NUP50 levels. Interestingly, nuclear NUP50 immunoreactivity was decreased in end stage transgenic mice expressing *Sod1*^G86R^, a severe ALS mouse model, as well as in mice expressing a truncated FUS protein (*Fus^ΔNLS/+^*), which display mild, late onset motor neuron degeneration (**Figure 5a-d**).Strikingly, some motor neurons of *Fus^ΔNLS/+^* mice displayed abnormal NUP50 immunoreactivity characterized by a perinuclear ring (**Figure 5b**). We then investigated the levels of NUP50 in hiPSC-derived motoneurons. In order to cover a large portion of the heterogeneous spectrum of ALS cases, we considered two patients harbouring mutations within the *FUS* gene, two with *TARDBP* pathogenic mutations, and two displaying the pathogenic HRE in the *C9ORF72* gene. After eight weeks in culture, we found that the nuclear levels of NUP50 were significantly lower in motoneurons differentiated from all ALS-related hiPSC lines, when compared to three healthy controls (**Figure 5c-d**). These results show that loss of nuclear NUP50 is a common event downstream of ALS mutations.

### NUP50 depletion leads to neuronal death and cytoplasmic nucleoporin inclusions

We then asked whether reducing NUP50 expression could have detrimental effects on neurons. Knockdown of *Nup50* through siRNA decreased mRNA (**Supplementalfigure 11**) and protein (**Figure 6a-b**) levels of NUP50 in HT22 mouse hippocampal neurons, while not affecting levels of other nucleoporins or of Ran and RanGAP1, two proteins critical for nuclear pore function (**Figure 6a-b**). However, consistent with a function of NUP50 in nucleocytoplasmic transport, *Nup50* knockdown abrogated nuclear export of a fluorescent NLS-mCherry-NES reporter^46^ (**Supplementalfigure 12a-b**). Most intriguingly, HT22 neurons showed cytoplasmic inclusions of nuclear pore proteins, as detected using mAb414, an antibody staining multiple nucleoporins (**Figure 6c-d**) and of Ran-GAP1 (**Supplementalfigure 12c-d**). Relevant to ALS, *Nup50* knockdown triggered also p62 positive inclusions, but not ubiquitin inclusions or stress granules (**Figure 6e-f** and **Supplementalfigure 12e-h**). Last, *Nup50* knockdown increased neuronal death in HT22 neurons (**Figure 6g**) and in primary cortical neurons (**Figure 6h**). Thus, loss of NUP50 compromises nuclear pore function and neuronal survival in cultured neurons.

### NUP50 depletion is sufficient to trigger motor phenotype *in vivo*

To further substantiate a pathogenic effect of reduced NUP50 levels in ALS, we knocked down *Drosophila Nup50*. We first evaluated the level of knock-down induced by a transgenic Nup50-RNAi line with no predicted off-targets, by ubiquitously (*actin5C-GAL4*) expressing Nup50-RNAi in third instar larvae and evaluating Nup50 transcript levels by qPCR. Nup50-RNAi expression reduced Nup50 transcript levels by 77% (**Figure 7a**). Next, we evaluated the effect of *Nup50* knock-down on motor performance. We utilized an automated negative geotaxis climbing assay^47^ to evaluate motor performance of 1- and 7-day-old adult flies. Whereas selective *Nup50* knock-down in motor neurons (*OK371-GAL4*) did not significantly affect motor capacities at 1 day of age, 7-day-old flies displayed a significant motor deficit (**Figure 7b-c**), indicative of an age-dependent progressive motor phenotype. Finally, we evaluated neuromuscular junction (NMJ) morphology in third instar larvae by measuring the length of the NMJs on distal muscle N°8, as shortening of NMJ length is frequently observed in *Drosophila* models of (motor) neurodegenerative diseases^47, 48^. Motor neuron-specific *Nup50* knock-down significantly reduced NMJ length on muscle 8 (**Figure 7d-f**), indicating a length-dependent NMJ phenotype.

To determine the consequences of NUP50 loss of function in a vertebrate model, we performed *nup50* knockdown by antisense-mediated oligonucleotides with the morpholino moiety in zebrafish embryos. *NUP50* has a single zebrafish orthologue (*nup50*; NM_201580.2) with 54% target identity to the human gene. We targeted the initial AUG of *nup50* by designing a specific antisense oligonucleotide morpholino (AMO). We did not observe any significant developmental deficits or non-specific toxicity, as shown at 48 hours post fertilization (hpf) (**Figure 7f-g**). However, only upon knockdown of *nup50*, we observed reduced swimming bouts in 50 hpf embryos (**Figure 7h**); and analysis of the touch-evoked escape response (TEER) showed that *nup50* knockdown by AMO led to impaired locomotion with embryos displaying significantly reduced velocity (**Figure 7i**). Importantly, we observed a significant reduction of zebrafish embryos displaying motor features upon co-expression of *nup50* knockdown alongside NUP50 human cDNA (hDNA) as compared to *nup50* AMO (**Figure 7g**). Similarly, analysis of the velocity from the TEER swimming bouts of *nup50* AMO + NUP50 hDNA showed no difference with the control conditions (**Figure 7i**).

We then analyzed motor neuron axonal projections using the *Hb9:GFP* transgenic line. We observed that motor neurons display abnormal axonal branching with reduced length upon *nup50* knockdown as compared to control conditions (**Figure 7j-k**). Significantly, co-expression of NUP50 human DNA alongside nup50 knockdown also rescued the deficits observed in axonal projections from spinal motor neurons, thus confirming the specificity of the phenotype of nup50 AMO embryos. (**Figure 7j-k**). These results further indicate that NUP50 loss of function is associated with motor deficits and specific alterations of the motor neuron axonal projections.

## Discussion

Our current study provides three major results in our understanding of ALS heritability. First, we provide genome wide evidence of the importance of variants affecting splicing in ALS heritability. Second, we identify a number of new candidate genes for ALS using TWAS. Third, we provide the first genetic evidence linking the core components of the nuclear pore to ALS.

We have shown that annotations constructed using fine-mapped posterior probabilities for sQTLs are enriched for ALS disease heritability. Independently, our PRS analysis show that fine-mapped sQTLs variants are significantly associated with ALS risk. What could be the relevance of these genetic results for ALS disease pathogenesis? It is noteworthy that ALS is characterized by the occurrence of phospho-TDP-43 aggregates, and concomitant loss of TDP-43 function. A major function of TDP-43 is to modulate splicing, and specifically, inclusion of cryptic exons. Recent work has shown that loss of TDP-43 leads to inclusion of cryptic exons in *STMN2*^49, 50^ or *UNC13A* mRNAs, leading to decreased levels of the encoded proteins that are critical for neuronal function. Recently, the risk allele of *UNC13A*, a major ALS GWAS signal, was found to synergize with loss of TDP-43 function to include a cryptic exon and decrease UNC13A levels^12, 13^. It is thus possible that the ALS heritability enrichment in sQTLs reflects a number of possible cryptic exons whose inclusion might be potentiated by TDP-43 aggregation. In this respect, two of our findings reinforce this hypothesis. First, we show that ALS heritability is enriched in TDP-43 and FUS binding sites, suggesting that splicing dysregulation mediated by these two proteins contributes to ALS risk. Second, our TWAS results identify decreased levels of *PTBP2* expression as a risk factor for ALS, and PTPB2 was shown to repress cryptic exons specifically in differentiated neurons^34^. Thus, our results are consistent with the notion that ALS heritability lies in part in altered TDP-43 and FUS splicing events. Further research should focus on the possible relationships between identified sQTLs and TDP-43/FUS dysfunction.

Another important conclusion from our analysis of heritability is that genetic variants associated with ALS are located mostly in genes expressed in neurons, in particular in inhibitory neurons. These results are consistent with the recent analysis of Van Rheenen and collaborators that showed enrichment in neuronally expressed genes using single cell RNA sequencing databases^7^. Here, using single cell ATACseq profiles, we observed similar enrichment in excitatory neurons, but also a significant association with inhibitory neurons. Importantly, both of our studies suggest a lack of a major contribution of microglia or astrocyte biology to ALS genetic risk, while we were able to correctly identify microglia expressed genes as contributing to AD. Of note, motor neurons or cholinergic neurons are currently not included in the datasets that we or Van Rheenen and colleagues used, and thus their involvement could not be studied. A potential role of inhibitory neurons, as our data suggest, is consistent with recent work in animal models. Indeed, structural, transcriptional or electrophysiological alterations in inhibitory neurons have been documented in mutant SOD1 mice^51–54^, TDP-43 transgenic mice^55^, TDP-43 knock-in mice ^56^, and recently in *Fus* knock-in mice ^57^. Furthermore, rescuing cortical inhibitory interneurons in mutant SOD1 mice appeared protective ^51^. Our results suggest a broader involvement of inhibitory neurons in ALS risk, which might open up additional therapeutic avenues.

In this study, we also identify several novel gene associations with ALS using brain specific TWAS. Interestingly, we were able to replicate previous gene associations that did not pass genome-wide significance for GWAS. This is in particular the case for *CX3CR1,* which has been associated with ALS^58–60^ but was not highlighted in previous GWAS, and for *ATXN3*, *MYO19 or SLC9A8*, which were also identified in previous TWAS^61, 62^. That our TWAS was able to pinpoint previously identified gene associations is strongly arguing for their relevance. In addition, a large fraction of the newly associated genes fall into functional categories that are known to be involved in ALS. For instance, a number of genes related to mitochondrial respiratory chain (*ATP5H, ATP5PD, BCS1L*) or mitochondrial lipid metabolism (*PGID, PGS1*) or mobility (*MYO19*). This echoes previous findings showing mutations in genes critical for mitochondrial function in ALS and FTD patients, such as *CHCHD10*^63^, and prominent mitochondrial dysfunction in cells carrying mutations in major ALS genes^64–66^. In addition, the TWAS identified candidates with immediate obvious relevance to known ALS pathogenic processes, including *PTBP2*, which shares with TDP-43 a function in repressing cryptic exons, and *NUP50*, encoding a nuclear pore protein, which we selected for further functional validation.

We focused on the *NUP50* locus for several reasons. First, its association with ALS was previously unknown, and reached significance in TWAS, but not in GWAS. Second, the function of NUP50 in the nuclear pore appeared highly relevant to ALS. Indeed, previous evidence linking genetically the nuclear pore to ALS were limited, and, to our knowledge, only mutations in *GLE1,* encoding a protein involved in mRNA export and translation and associated to the nuclear pore, have been found in ALS patients^67^. Third, the association in TWAS was specific to brain, and not found in blood. Interestingly, NUP50 has been previously associated to ALS pathogenic processes, although with divergent results. Loss of *Nup50* enhanced the toxicity of GGGGCC repeats in *Drosophila* ^14^, but suppressed the toxicity of a PR25 di-peptide repeat^17^ or mutant TDP-43^21, 68^. NUP50 appeared to be the only nucleoporin tested to interact more avidly with full length TDP-43 than with its C-terminal fragment^21^ and suggests a complex functional relationship between NUP50 and TDP-43, possibly linked to NUP50 complex function in the nuclear pore and gene expression^69, 70^ or DNA damage^71, 72^. Of further interest, NUP50 was recently observed to be decreased in cortical neurons of *C9ORF72* HRE patients, as well as in motor neurons differentiated from *C9ORF72* HRE patients^45^. Our own results confirmed and extended those results by showing that NUP50 loss occurs in nuclei of motor neurons differentiated from *C9ORF72* but also from *TARDBP* and *FUS* patients or *SOD1* and *Fus* mouse models. Altogether, this evidence pointed out that the *NUP50* locus is of specific interest in our TWAS results. Confirming *NUP50* relevance, we observed suggestive enrichment in rare variants through burden analysis, although these variants remained rare and larger WES studies are required to replicate this signal. It remains uncertain whether the TWAS signal observed is solely due to the burden of *NUP50* variants identified, or if other mechanisms, such as splicing alterations at the *NUP50* locus also contribute to the observed association. Importantly, we identified a number of *NUP50* variants in ALS patients, predicted to be highly damaging, and the two frameshift mutations may lead to haplo-insufficiency. Analysis of cells or tissues from two of these patients confirmed that ALS associated *NUP50* gene variants may lead to haploinsufficiency.

What could be the consequences of *NUP50* loss? Interestingly, *Nup50* knock out mice die *in utero* due to severe neural tube defects ^73^, suggesting a critical function of NUP50 in the central nervous system. Consistent with this hypothesis, knockdown of *Nup50* in primary neurons or in HT22 cell lines increased neuronal death. We also observed cytoplasmic inclusions of nuclear pore components, as well as of RanGAP1,a key protein regulating nucleocytoplasmic shuttling function, which recapitulates previous observations in ALS patients^15, 21^. These cytoplasmic nuclear pore inclusions were analogous to the inclusions recently observed upon *FXR1* downregulation ^74^. Hence, further research is needed to determine their origin and their potential relevance to ALS, especially as these cytoplasmic nucleoporin inclusions co-occur with p62 inclusions also found in ALS patients, in particular *C9ORF72* HRE patients^75–77^. Consistent with our results, defects in nucleocytoplasmic transport, similar to what triggered by loss of *NUP50* expression, have been recently positioned upstream to autophagy/lysosome defects (including p62 accumulation) in *C9ORF72* ALS^78^. To clarify the *in vivo* relevance of reduced *NUP50* expression, we further generated and characterized two animal models with partial loss of NUP50 function. In Drosophila, knock-down of *Nup50* in motor neurons led to a mild but significant loss of motor function accompanied by decreased neuromuscular junction size. Our results are in accordance with previous studies, in which *Nup50* loss of function had no effect on motor neuron development in *Drosophila* ^21^. Indeed, we did not observe motor defects in 1 day old adult flies, but the defect appeared later in life, consistent with ALS being an adult onset disease. Similarly to *Drosophila* experiments, *nup50* knockdown in zebrafish led to reduction of evoked swimming bouts, without any apparent developmental deficits. Importantly, this locomotor phenotype was associated with abnormal motor neurons morphology characterized by a shortening of their axonal projections. Both these motor features were rescued upon co-expression of NUP50 human cDNA. Altogether, our results show that loss of function of NUP50 is toxic to motor neurons, and sufficient to lead to motor neuron disease. This is consistent with *NUP50* haploinsufficient mutations being causative of ALS.

Summarizing, our current studies show that ALS is mostly associated with variants affecting splicing and protein expression in neuronally expressed genes, and a number of new loci associated to ALS identified here. We further validate these genetic discoveries by validating the functional effect of reduced *NUP50* expression as a contributing factor in ALS, thus providing a genetic link between ALS and nuclear pore defects.

## Methods

### Genetics

#### GWAS summary association statistics

We used the summary association statistics from the latest GWAS of ALS^7^. Also summary association statistics for SCZ and ASD were obtained from the PGC consortium (https://www.med.unc.edu/pgc/). AD and PD summary association statistics were obtained from the latest GWAS^79, 80^.

#### Individual level genotype data

We obtained available summary statistics from the largest ALS GWAS to date. We obtained individual genotype level data that contains 10067 ALS cases (accession number phs000101.v5.p1). For the control cohort, we used genotype data obtained from the database of Genotypes and Phenotypes (dbGaP), (a) the Cardiovascular Health Study (n=3802), (b) Genetic Epidemiology of Refractive Error in the KORA Study (n=1863), (c) Genetics of Schizophrenia in a Ashkenazi Jewish Case-Control Cohort (n=2052), (d) Multi-Ethnic Study of Atherosclerosis (MESA) Cohort (n=3821), PAGE: The Charles Bronfman Institute for Personalized Medicine (IPM) BioMe Biobank (n= 3700).

#### Quality control and imputation

For each cohort, SNPs were first annotated according to dbSNP150 and mapped to the hg19 reference genome. All multi-allelic and palindromic (A/T or C/G) SNPs were excluded. Low quality SNPs and genotyped individuals were excluded using PLINK 1.9 (--geno 0.02 and --mind 0.1). The following filter criteria were applied: MAF > 0.01, SNP genotyping rate > 0.98, Deviation from Hardy-Weinberg disequilibrium in controls P > 1 × 10-5. Then, more stringent QC thresholds were applied to exclude individuals: individual missingness > 0.02, inbreeding coefficient |F| > 0.2, mismatches between genetic and reported gender, and missing phenotypes (PLINK --mind 0.02, --het, --check-sex). Duplicate individuals were removed (PI_HAT > 0.8). Population structure was assessed by projecting 1000G principal components (PCs) and outliers from the European ancestries population were removed (> 4 SD on PC1-4) (**Supplemental figure 13).** Finally, samples in common between the individual genotype data and van Rheenen’s study were identified using the checksum program id_geno_checksum and were removed from our analyses. Finally, 7584 cases and 11280 controls were used for imputation. In each cohort, between 278,390 and 635,011 SNPs passed QC and were included for imputation.

#### Post Imputation quality control

Cohorts were then imputed using the HRC reference panel (r.1.1 2016) on the Michigan Imputation Server^81^. Data was phased using Eagle 2.3. Post-imputation variant-level quality control included removing all monomorphic SNPs and multi-allelic SNPs from each cohort. SNPs with MAF < 1% in the HRC imputation panel were excluded. Subsequently, INFO scores were calculated for each cohort based on dosage information using SNPTEST v2.5.4-beta3. Within each cohort, SNPs with an INFO-score < 0.3 and those deviating from Hardy-Weinberg equilibrium at P < 1 × 10-5 in control subjects were removed. All cohorts were combined including SNPs that passed quality control in every cohorts yielding 7,331,489 autosomal SNPs.

#### Transcriptomics panels for TWAS

The following panels were used in our study: (1) BrainSeq DLPFC and HIPPO RNA-seq data generation and processing were previously described^82^. Ready-to-analyze genotype were accessed through dbGaP (phs000979.v3.p2). (2) CMC RNA-seq data: generation and processing were previously described^83^. Briefly, DLPFC (Brodmann areas 9/46) was dissected from post-mortem brains of 258 individuals with schizophrenia and 279 control subjects. These individuals were of diverse ancestry, had no AD or PD neuropathology, had no acute neurological insults (anoxia, stroke, or traumatic brain injury) immediately before death, and were not on ventilators near the time of death. Total RNA was isolated from homogenized tissue and ribosomal RNA depleted. One hundred base pair paired-end reads were obtained using an Illumina^_^; HiSeq 2500, and mapped using TopHat. Genotyping was performed using the Illumina Infinium HumanOmniExpressExome-8 v1.1b chip. (3). Data is provided for the Mayo RNAseq Study, with whole transcriptome for 276 Temporal cortex (TCX) samples from 312 North American Caucasian subjects with neuropathological diagnosis of AD, progressive supranuclear palsy (PSP), pathologic aging (PA) or elderly controls (CON) without neurodegenerative diseases ^84^. Within this cohort, all AD and PSP subjects were from the Mayo Clinic Brain Bank (MCBB), and all PA subjects were obtained from the Banner Sun Health Research Institute (Banner). Mayo RNAseq samples samples underwent 101 bp, paired-end sequencing. Genotypes were obtained for all samples in the Mayo RNAseq Study utilizing Illumina’s Human Omni 2.5 + Exome array. The sample QC included removal of samples with a mismatch between the recorded sex and the sex deduced by evaluating SNPs on the X chromosome, call rates < 98%, heterozygosity rate > 3SD from the mean, identity by descent PI_HAT > 0.04, while keeping the subject with the better call rate in each related pair, and removing population outliers using Eigensoft v.6.0.1 (for details see the biospecimen metadata). The SNP QC included removal of SNPs with genotyping call rate < 98%, minor allele frequency < 0.02, Hardy-Weinberg disequilibrium p < 3.4×10-8 in controls, duplicate variants and multiallelic SNPs. After sample and SNP QC, genotype data was retained for 303 subjects. For all transcriptome datasets used in this study see **Supplemental table 11.**

#### TWAS studies

TWAS is a powerful strategy that integrates SNP-expression correlation (*cis*-SNP effect sizes), GWAS summary statistics and LD reference panels to assess the association between the *cis*-genetic component of expression and GWAS. TWAS can leverage large-scale RNA-seq and/or methylation data to impute tissue-specific genetic expression levels from genotypes (or summary statistics) in larger samples, which can be tested to identify potentially novel associated genes^31^.. We used the FUSION tool to perform TWAS for each transcriptome reference panel. The first step in FUSION is to estimate the heritability of each feature (gene expression or intron usage) using a robust version of GCTA-GREML^85^, which generates heritability estimates per feature as well as the likelihood ratio tests *p* value. Only genes or intron usage that were significant for heritability estimates at a Bonferroni-corrected *p* < 0.05 were retained for further analysis. The expression or intron usage predictive weights were computed by four different models implemented in the FUSION framework: best linear unbiased prediction, LASSO, Elastic Net, and top SNPs. A cross-validation for each of the desired models are then performed. The model with the best cross-validation prediction accuracy are used for predicting expression or intron usage into the GWAS. The imputed gene expression or intron usage are then used to correlate to ALS GWAS summary statistics to perform TWAS and identify significant associations. To account for multiple hypotheses, we applied an FDR of 5% within each expression and splicing reference panel (see “Transcriptomics panels for TWAS”) that was used.

#### Joint and conditional analysis

Joint and conditional analyses of each locus with multiple TWAS association signal were performed using the summary statistic-based method^86^. This approach requires TWAS association statistics and a correlation matrix to evaluate the joint/conditional model. The correlation matrix was estimated by predicting the *cis*-genetic component of expression (or intron usage) for each TWAS gene/intron cluster into the 1000 Genomes genotypes. Then, Pearson’s correlations were calculated across all pairs of genes/intron cluster and between all gene/SNP pairs. The FUSION tool (see URLs) was used to perform the joint and conditional analyses and generate regional scatterplots.

#### Colocalization

We used coloc 2.3-1 ^87^ to colocalize the ALS association signal at TWAS loci with QTL signals. For each locus, we examined all SNPs available in both datasets within 500 Mb of the SNP identified in TWAS as the top QTL SNP, and ran coloc.abf with default parameters and priors. We called the signals colocalized when (coloc H3 + H4 ≥ 0.8 and H4/H3 ≥ 2).

#### Splicing QTLs mapping (sQTLs)

For the BrainSeq phase 1, 2, Mayo and CMC cohorts, we used Leafcutter^32^ to obtain intron excision ratios, which is the proportion of intron defining reads to the total number of reads from the intron cluster it belongs to. We used the alignments from STAR as an input to LeafCutter. The intron excision ratios were standardized across individuals for each intron and quantile normalized across introns. The normalized intron excision ratios were used as our phenotype matrix. To map sQTLs, we used linear regression (implemented in fastQTL^86^ to test for associations between SNP dosages (minor allele frequency (MAF) > 0.01) within 100 kb of intron clusters and the rows of our phenotype matrix that correspond to the intron excision ratio within each intron cluster. We used the first three principal components of the genotype matrix to account for the effect of ancestry plus the first 15 principal components of the phenotype matrix to regress out the effect of known and hidden confounding factors. An adaptive permutation scheme^86^ (implemented in fastQTL) was used to estimate the number of sQTLs for any given FDR. An empirical *p* value for the most significant QTL for each intron cluster was calculated.

#### Fixed-effect meta-analysis of brain QTL effect sizes (FE-meta-Brain)

Meta-analysis was performed via fixed-effect model using an adaptation of the metareg function in the gap package in R. Given a set of effect sizes for SNP *i* (*β*_1_, *β*_2_, … *β_s_*) for *s* studies, where *β_j_* is the eQTL effect size for study *j*, we used fixed-effect meta-analysis (FE-meta-Brain) to compute inverse-variance weighted meta-analysis *z*-scores *z*_FE_ as follows:

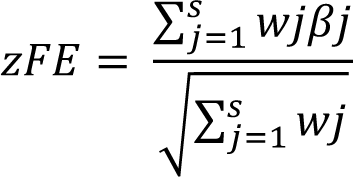

Where 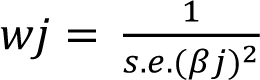 and s.e.(*β_j_*) is the standard error of *β_j_*. We note that equation (1) is equivalent to computing a weighted average of *z*-scores^88^. After filtering, for low expressed genes, we used ∼3 million eQTLs in 13965 genes, ∼3.3 million sQTLs in 15463 genes, ∼660.000 mQTLs in 46656 CpG sites and ∼1.5 millions pQTLs in 8167 proteins.

### Brain QTLs-based annotations

We construct the annotations for any given Brain QTL datasets using the observed marginal association statistics as previously described^88^. Briefly, each annotation is a vector that assigns a value to each SNP. Given **a** indicate our annotation for one QTL dataset where ***a****_j_* indicates the value assigned to SNP *j*. For continuous probabilistic annotations (MaxCPP), 0 ≤ ***a****_j_* ≤ 1. Let *S* = (***s***_1_, ***s***_2_, …***s****_g_*) indicate an *m* × *g* matrix of the observed marginal association statistics obtained for each QTL dataset, where *m* is the number of SNPs and *g* is the number of eGenes (for example, genes that have at least one significant *cis*-eQTLs). Let **s***_i_* be the vector of marginal association statistics of gene *i* for all cis variants. Utilizing **s***_i_* and the LD structure, we can compute the CPP for each variant. CPP is the probability that a variant is causal. Given *α_ji_* the posterior probability that SNP *j* is causal for gene *i*. We obtained the CPP values from CAVIAR^24^. In addition to the CPP values, CAVIAR provides a 95% credible set that contains all of the causal variants with a probability of at least 95%. We constructed the MaxCPP annotation for SNP *j* by computing the maximum value of CPP over all genes for which SNP *j* is in the 95% credible set of gene *i*. More formally, we have: *a_j_* = max*_i_α_ji_* where the maximum is over genes *i* with *θ_ji_* = 1.

### Estimating the RBP dysregulation GWAS effect sizes

We use the previously published statistical framework of stratified LD score regression^79^ to estimate the RBP dysregulation effect sizes for each examined disease GWAS. From the summary statistics of a GWAS, we can write the expected χ2 value for SNP j as:

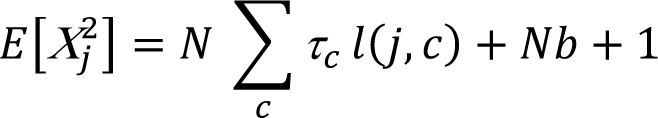

where N is sample size and the annotation specific “LD score” l(j, c), representing annotation (c)’s cumulative effects tagged by the SNP j, can be written as:

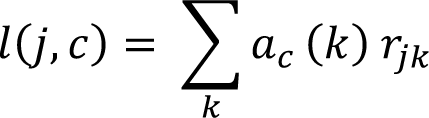

where a_c_(k) is the annotation value at SNP k (e.g RBP dysregulation level or molecular QTLs), r_jk_ is the correlation between SNP j and k in the reference panel (selected to best match the GWAS cohort), and b measuring the confounding bias^79^. Lastly, τ_c_ and the final standardized form τ_c_* normalized by the total SNP-based heritability and s.d. of an annotation – represents the estimated effect size of the annotation^79^.

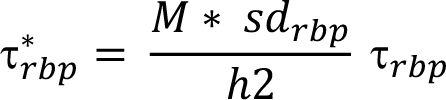

For RBP dysregulation annotations, τ* represents the per-SNP heritability associated with a standard deviation increase of variant RBP effect (sd_rbp_). We fit τ*_rbp_ by conditioning on a baseline annotation. The final reported RBP effect sizes (τ*_rbp_) were jointly fit, iteratively for each RBP, with all baseline annotations. We have made the annotation for each of the 125 RBPs available for public use (link).

### LD-score regression and partitioned heritability for ALS

After obtaining the MaxCPP annotations, we ran S-LDSC^89^ to generate the LD score of each variant in each annotation using the same procedure described previously ^24^. Regression SNPs, which are used by S-LDSC to estimate *τ* from marginal association statistics, were obtained from the HapMap Project phase^90^. Using the LD score for each annotation and the marginal statistics obtained from the trait phenotypes (GWAS-ALS), we computed the enrichment, the coefficient *τ* and calculated the p-value for each annotation conditional on the baseline model v1.1 and all MaxCPP annotation^91^. For cell-type enrichment analyses^25^ we used human-derived single-cell ATAC sequencing data on major brain cell-types (GSE147672).. We then confirmed our functional enrichment using PRS calculation in functional categories. Disease-relevant gene-sets annotation (MaxCPP-ExAC) were constructed as previously described^88^. Briefly, eQTLs and sQTLs annotation were restricted to the 3,230 genes that are strongly depleted of protein-truncating variants^28^.

### Polygenic risk score

We used a three-stage study design to identify molecular QTLs relevant to ALS risk as described previously^30^. We separated the available ALS genomic data into three independent datasets for analyses. The reference dataset consisted of summary statistics from a previous published GWAS involving 12,577 cases and 23,475 controls of European ancestry (publicly available from databrowser.projectmine.com). We used the summary statistics from this reference dataset to define risk allele weights for constructing polygenic risk scores within the 5 MaxCPP annotations built for each molecular trait. The remaining data consisted of individual-level genotype and phenotype data from 7,584 ALS cases and 11,280 control subjects of European ancestry recently published ^92^. We split these data in a 70%-to-30% ratio into a “training dataset” containing 5,226 cases and 7,866 control subjects, and a “replication dataset” consisting of 2,298 patients and 3,314 controls subjects. We used the regression model generated from the reference data to construct and test polygenic risk scores within the training data. The replication dataset was used to validate our training data findings. There was no sample overlap between the reference, training, or replication datasets. The target dataset consisted of individual-level dosage and phenotype.

Polygenic risk scores were calculated based on the weighted allele dosages as implemented in PRSIce2. For the training dataset, 1,000 permutations were used to generate empirical p-value estimates for each GWAS-derived p-value. Each permutation test in the training dataset provided a Nagelkerke’s pseudo R2 value after adjusting for an estimated ALS prevalence of 5 per 100,000. Sex and eigenvectors one to twenty were included as covariates in the model. The PRSset function of PRSice2 was used to include 5 different sets of SNPs corresponding to the five molecular traits after filtering for a CPP value > 0.9. We selected variants in each annotation that have a CPP value above 0.9 yielding ∼6000 eQTLs, ∼8000 hQTLs, 16000 mQTL, ∼4000 pQTLs and ∼7000 sQTLs. We applied the default clumping parameters outlined in the PRSice2 software package^93^ (version 2.1.1, R2 = 0.1, and a 250 kb window, GWAS p-value = 0.05). This clumping process yielded a total of 49,027 variants that were then used for polygenic risk score analysis. The number of variant used in each annotation is shown in between brackets in **Figure 1d**.

Polygenic risk scores were then tested in the replication phase using the --score command implemented in PLINK v1.9^94^. Polygenic risk scores were calculated, incorporating the risk variants from the molecular QTLs annotation nominated in the discovery phase. All SNPs were weighted by the log-odds ratios obtained from the reference dataset with a greater weight given to alleles with higher risk estimates. Logistic regressions were performed to evaluate the association between the molecular QTLs polygenic risk score of interest with ALS as the outcome.

### Cell-type specific QTL risk score analysis

Single cell ATAC-seq peaks generated previously^25^ were used as an input to PRSice2. Briefly, annotation files for 12 different cell types were liftOver to hg19 genomic annotation. PRSet was run as described above with bed-files as input with default clumping procedure. The clumping process yielded a total of 58,182 variants that were then used for cell-type polygenic risk score analysis. We then intersected annotated SNPs as being a significant molecular QTL with a significant associated cell-type. With this new set of SNPs, polygenic risk scores were then tested in the replication phase using the --score in PLINK1.9 as described above.

### Genic burden association analyses

To aggregate rare variants in a genic burden test framework we used the results of Datafreeze 2 from Project MinE as described in the ALS 2021 GWAS^7^. In short, a variety of variant filters was applied to allow for different genetic architectures of ALS associated variants per gene as was used previously^95, 96^. In summary, variants were annotated according to allele-frequency threshold (MAF < 0.01 or MAF < 0.005) and predicted variant impact (“missense”, “damaging”, “disruptive”). “Disruptive” variants were those variants classified as frame-shift, splice-site, exon loss, stop gained, start loss and transcription ablation. “Damaging” variants were missense variants predicted to be damaging by seven prediction algorithms (SIFT^97^, Polyphen-2^98^, LRT^99^, MutationTaster2^100^, Mutations Assessor^101^, and PROVEAN^102^). “Missense” variants are those missense variants that did not meet the “damaging” criteria. All combinations of allele frequency threshold and variant annotations were used to test the genic burden on a transcript level in a Firth logistic regression framework where burden was defined as the number of variants per individual. Sex and the first 20 principal components were included as covariates. All ENSEMBL protein coding transcripts for which at least five individuals had a non-zero burden were included in the analysis.

### Patients

#### Ethics

For the French cohorts, protocols for genetic analyses and gene expression analysis in patient lymphoblasts were approved by the Medical Research Ethics Committee of “Assistance Publique Hôpitaux de Paris” (#A75). All FALS and ySALS patients signed a consent form for the genetic research. Autopsied patients were enrolled in the NeuroCEB brain donation program declared to the Ministry of Research and Universities, as requested by French Authorities (#AC-2013-1887). An explicit consent was signed by the patient himself, or by the next of kin, in the name of the patient, in accordance with the French Bioethical Laws. This document includes consent for genetic analyses. The diagnosis of ALS and FTD was based on published criteria as previously described^103^.

All procedures with human materials were in accordance with the ethical committee of Ulm University (Nr.19/12 and Nr. 135/20) and in compliance with the guidelines of the Federal Government of Germany. All participants or their next of kin gave informed consent for the study. The use of human materials was compliance with the Declaration of Helsinki concerning Ethical Principles for Medical Research Involving Human Subjects, and experiments were performed according to the principles set out in the Department of Health and Human Services Belmont Report. Tissue from ALS patients and controls used in western blotting and immunofluorescence were obtained from the Neurology Brain Bank at Ulm University, Germany. Tissue from Patient 2 was provided by the Kantonsspital St. Gallen (KSSG), Switzerland.

#### Genetic analyses

Whole Exome Sequencing (WES) analyses were performed on French patient cohorts (including 200 family members with 150 index cases and 180 sporadic cases with 100 patients with ALS and 80 autopsied cases) as previously described ^43^. These patient cohorts were devoid of mutation in the 4 major ALS genes (including repeat expansions in *C9orf72* and/or mutation in *SOD1, TARDBP* and *FUS*). These exome databases were interrogated to select heterozygous or homozygous qualifying variants in coding or non coding region of *NUP50* gene with a minor allele frequency (MAF) threshold <0.005% in gnomAD database for heterozygous variants and ≤1% for homozygous variants ^42^.

Any patient with a *NUP50* variant was further interrogated for a list of 35 rare ALS related genes (including *ALS2, ANG, ANXA11, ATP13A2, CCNF, CHCHD10, CHMP2B, DAO, DCTN1, DNAJC7, ERLIN2, FIG4, GLE1, GLT8D1, HNRNPA1, HNRNPA2B1, KIF5A, MAPT, MATR3, NEK1, OPTN, PFN1, PRPH, SETX, SIGMAR1, SPG11, SQSTM1, SS18L1, TAF15, TBK1, TIA1, TUBA4A, UBQLN2, VAPB* and *VCP*) to select variants with a MAF <0.005% in gnomAD database. The *NUP50* frameshift variant was validated using Sanger analysis with BigDye chemistry as recommended by the supplier (Applied Biosystems).

#### Lymphoblast cultures

Lymphoblastoid cell lines from the French ALS patient carrying the NUP50 frameshift variant were established by Epstein Barr virus transformation of peripheral blood mononuclear cells. Lymphoblasts from four healthy age-matched individuals were used as control. Lymphoblasts were grown in RPMI 1640 supplemented with 10% fetal bovine serum, 50 U/ml penicillin and 50 mg/ml streptomycin (Life Technologies) renewed twice a week.

#### Western blotting of lymphoblast extracts

Lymphoblasts cells were homogenized in lysis buffer (50 mm Tris, pH 7.4, 150 mm NaCl, 1 mm EDTA, pH 8.0, and 1% Triton X-100) containing protease and phosphatase inhibitors (Sigma-Aldrich) and protein concentration was quantitated using the BCA protein assay kit (Pierce). Fifteen micrograms of proteins were loaded into a gradient 4–20% SDS-PAGE gel (Bio-Rad, 5678094) and transferred on a 0.45 µm nitrocellulose membrane (Bio-Rad) using a semi-dry Transblot Turbo system (Bio-Rad). Membranes were saturated with 10% nonfat milk in PBS and then probed with the anti-NUP50 (Abcam, ab137092, 1:1000) primary antibody diluted in 3% nonfat milk in PBS. Blots were washed and incubated with anti-Rabbit secondary antibody conjugated with HRP (P.A.R.I.S, BI2407, 1:5000) for 2 hours. Membranes were washed several times and analyzed with chemiluminescence using ECL Lumina Forte (Millipore, WBLUF0500) using the Chemidoc XRS Imager (Bio-Rad). Total proteins were detected with a stain-free gel capacity and normalized.

#### RT-qPCR of lymphoblast RNA

1 µg of RNA was reverse transcribed with iScript™ reverse transcription (Biorad, 1708841). Quantitative polymerase chain reaction was performed using Sso Advanced Universal SYBR Green Supermix (Bio-Rad) and quantified with Bio-Rad software. Gene expression was normalized by calculating a normalization factor using *ACTIN*, *TBP* and *POL2* genes according to GeNorm software ^104^

Primer sequences were as follows:

*ACTIN*: 5’- GGGCATGGGTCAGAAGGATT-3’, 5’-TCGATGGGGTACTTCAGGGT-3’

*TBP*: 5’-TGACCCAGGGTGCCATGA-3’, 5’-GGGTCAGTCCAGTGCCATAA-3’

*POL2*: 5’-CGGCCTGTCATGGGTATTGT-3’, 5’-TTCATCACTTCACCCCGCTC-3’

*NUP50*: 5’- CATTCTCCATGGCCAGTGAG-3’, 5’- AAAAGCGTCCTCCTCCAGAA-3’

### RT-PCR from post-mortem tissue

RNA from natively frozen *post-mortem* motor cortex tissue was isolated using RNAzol RT (Sigma-Aldrich #R4533) combined with the Direct-zol RNA Miniprep Kit (Zymo Research #R2050) according to the instructions of the manufacturer. Reverse transcription was carried out with the iScript cDNA Synthesis Kit (Bio-Rad #1708890). NUP50 cDNA was amplified using oligonucleotides spanning exons 5 to 8 (5’-CAAAGATACTACCCAGAGTAAACC-3’ and 5’- TGGTTTTTACCCGAATCAACATGG-3’) resulting in a PCR product of 443 bp for the wildtype. As a control, a 200 bp fragment of GAPDH mRNA was amplified from the same samples using oligonucleotides 5’-GTCAAGCTCATTTCCTGGTATGAC-3’ and 5’- TGTGAGGAGGGGAGATTCAGT-3’.

### Allele-specific sequencing of NUP50 (patient 2)

Presence of rs773541780 in one *NUP50* allele of patient 2 allowed design of allele-specific oligonucleotides (5’-GTTACCGAAGTAAAAGAAGAAGACGC-3‘ and 5‘- AGGTTTTAAATGCAGAGTACCTATGC-3’) for amplification of a 2145 bp fragment (without rs773541780) covering the nearsplice mutation c.1086-6C>T. The fragment was used as a template for standard Sanger sequencing.

### Human post-mortem histology

Human motor cortex from aged controls and disease patients were sampled and subsequently frozen on dry ice (during the autopsy) and stored at −80°C. Tissue blocks from motor cortical area were than fixed in a 4% formaldehyde solution over weekend and embedded in paraffin. Paraffin blocks were cut through a microtome (Slee Medical GmbH, Mainz, Germany) to obtain 7 µm thickness sections. The sections were subsequently used for immunofluorescence staining.

### Immunofluorescence of paraffin sections

Thin paraffin sections were completely deparaffinized and rehydrated by immerging the slides through the following solutions: (1) Xylene (three washes 5 min. each); (2) 100% Ethanol (two washes 10 min. each); (3) 96% Ethanol (two washes 5 min. each); (4) 70% Ethanol (two washes 5 min. each); (5) deionized Water (two washes for 5 min.). After deparaffinization and rehydration step the sections were treated with 30% H_2_O_2_ in deionized water for 20 min for background reduction. For heat induced antigen retrieval, the slides were covered with 10 mM Sodium Citrate buffer (pH 6.0) at a boiling temperature (100 °C) and put in a steamer for 25 min. Sections were than blocked in a solution of 5% BSA + 10% NDS with addition of 0.03% Triton X-100 and incubated for 2 hs at RT. After blocking, the sections were incubated with primary antibodies against NUP50 (rabbit, Proteintech, 20798-1-AP, 1:2500) and phospho-TDP-43 (rat, Millipore, MABN14, 1:800) for 48 hs in Tris buffer at +4 °C. Primary antibodies were visualized by incubating the sections with Alexa Fluor® 488 and 568 secondary antibodies (Invitrogen; Life technologies) in Tris buffer for 2 hs at RT in a light protected environment. Nuclei were detected by adding DAPI (ThermoFisher Scientific, P36934) to the secondary antibody cocktail at the concentration of 1:500. Sections were finally coverslipped with Vectashield® anti-fade mounting medium (Vector Laboratories). Pictures were acquired with a confocal microscope LSM-710 (Carl Zeiss confocal AG).

### Confocal microscopy and image processing

Confocal images were acquired as previously reported (Ouali Alami et al, 2018). Briefly, images were acquired in single tiles using an LSM-710 (Carl Zeiss AG) inverted microscope fitted with a 40X oil objective. Images were captured in a 12-bit format at a resolution of 1024×1024 pixels (x-y) and a theoretically optimal optical section thickness (z). Imaging parameters were set in order to obtain a minimal signal intensity for the immunostained antigen >150 (range 0-4095 in 12-bits images) while avoiding saturation. Imaging parameters were kept constant across imaging sessions and samples. All images were acquired within the grey matter of the motor cortex, in correspondence of Betz cells location (Fig..) and further processed and analysed by using Image J software.

### Image analysis

For the quantification of NUP50 protein expression in nuclear and perinuclear region of Betz cells, the images were imported using FIJI image analysis software. Confocal stacks (5-9 1µm-thick optical sections, 40x oil objective) were collapsed in a maximum-intensity projection.

Regions of interest (ROIs), corresponding to the area surrounding the nucleus were manually traced (using the DAPI staining as reference), and quantified; the integrated mean gray value was obtained. Fluorescence intensity was expressed in arbitrary units (a.u) corresponding to the grayscale value (in 12-bits images, ranging from 0 to 4095). The positive NUP50 nuclear and perinuclear staining was also quantified considering the positive area under threshold. The resulting ratio between the area occupied by NUP50 and the total nuclear and perinuclear area was plotted in a dot-plot graph. The same procedure was used for the quantification of p-TDP-43 staining in the cytoplasmic region of ALS cases. For quantitative analysis, a minimum of 5–8 artefact-free sections per case and from each a minimum of 4-5 MNs were taken into consideration.

## Statistical analysis

One-way ANOVA with Bonferroni correction for multiple comparisons was applied for the comparison of multiple groups. Statistical analysis was performed with Prism software (GraphPad 8). All values were expressed as mean ±SD unless otherwise indicated. Statistical significance was set at P<0.05 before multiple comparison correction.

### Histology in mouse models

#### Histological techniques

Mouse experiments were approved by local ethical committee from Strasbourg University (CREMEAS) under reference number 2016111716439395 and 25452. Mice were anesthetized with intraperitoneal injection of 100 mg/kg ketamine chlorhydrate (Imalgène 1000®, Merial) and 5mg/kg xylazine (Rompun 2%®, Bayer), and then transcardially perfused with cold PFA 4% in phosphate buffered saline (PBS). After dissection, spinal cords were post-fixed for 24 hours and then included in agar 4% and serial cuts of 40 µm thick were made using vibratome (Leica Biosystems, S2000).

#### Immunohistochemistry

Free-floating sections were pre-treated 30 minutes in citrate buffer 0.1M pH 6.0 at 80°C, rinsed with PBS 1X, then incubated 30mn with blocking solution (5% Horse Serum Albumin, 1% Triton×100 in PBS). Sections were incubated overnight at 4°C with primary antibodies diluted in PBS+0.1% triton×100. The following antibodies were used: rabbit anti-Nup50 antibody (Ab 137092 Abcam, 1/200) and goat anti-choline acetyl transferase (AB144P, Millipore, 1/100). After 3 rinses in PBS, sections were incubated for 1h at room temperature with Hoechst (Sigma, B2261, 1/50.000) and secondary antibody: Donkey anti-rabbit Alexa-488 (JacksonImmunoResearch, 711-547-003, 1:1000) and donkey anti-goat Alexa-594 (Invitrogen, A11058, 1:1000). Finally sections were subsequently washed with PBS (3 x 10 min) and mounted in Aqua/polymount (Polysciences, 18606).

#### Confocal microscopy and analyses

Z-Stack images (1µm optical section, frame resolution of 1024×1024 pixels) were acquired using a laser scanning microscope (confocal LSM 800 Zeiss) equipped with 40×oil objective (NA1.4) zoom x 0.5. Excitation rays are sequential diodes 488nm, 561nm, 405nm. Emission bandwidths are 400-500nm for Hoechst, 500-570nm for Alexa488 and 570-617nm for Alexa594. Each Stack of 10µm were aplane and analyze using ImageJ freeware. First, the user defined mean intensity of Nup50 in motoneurone nucleus (ChAT positive cells) for 6 spinal cords per FUS or WT genotype or 3 spinal cords per SOD or WT gentotype. These values are divided in 5 categories of pixels intensity (0-50, 51-100,101-150,151-200, 200-255) and used to calculate nuclear Nup50 levels.

#### Human iPSCs

The hiPSC lines used in this study are detailed in **Supplemental Table 12.** The lines generated at Ulm University have been previously published ^105, 106^ while the other lines have been commercially purchased. hiPSC were cultured using mTeSR1 medium (Stem Cell Technologies, 83850) on Matrigel^®^-coated (Corning, 354277) 6-well plates using at 37°C (5% CO_2_, 5% O_2_). When colonies reached 80% confluence, they were passaged in 1:6 split ratio after detachment using Dispase (Stem Cell Technologies, 07923).

#### Differentiation of hiPSC-derived MN

We differentiated MN from hiPSCs as previously described ^106^. Briefly, confluent hiPSC colonies were detached and transferred ultra-low attachment T25 flasks (Corning, CLS430639) for the formation of embryoid bodies (EBs) in hESC medium (DMEM/F12 + 20% knockout serum replacement + 1% NEAA + 1% β-mercaptoethanol + 1% antibiotic-antimycotic + SB-431542 10 µM + Dorsomorphin 1 µM + CHIR 99021 3 µM + Pumorphamine 1 µM + Ascorbic Acid 200ng/µL + cAMP 10 µM + 1% B27 + 0.5% N2). On the fourth day of culture in suspension, medium was switched to MN Medium (DMEM/F12 + 24 nM sodium selenite + 16 nM progesterone + 0.08 mg/mL apotransferrin + 0.02 mg/mL insulin + 7.72 *μ*g/mL putrescine + 1% NEAA, 1% antibiotic-antimycotic + 50 mg/mL heparin + 10 μg/mL of the neurotrophic factors BDNF, GDNF, and IGF-1, SB-431542 10 µM, Dorsomorphin 1 µM, CHIR 99021 3 µM, Pumorphamine 1 µM, Ascorbic Acid 200ng/µL, Retinoic Acid 1 µM, cAMP 1 µM, 1% B27, 0.5% N2). After 5 further days, EBs were dissociated into single cells with Accutase (Sigma Aldrich, A6964) and plated onto μ-Plates (Ibidi, 82406) pre-coated with Growth Factor Reduced Matrigel (Corning, 354230).

#### Immunocytochemistry

Immunostainings were performed as previously described^106^. Cells were fixed with 4% paraformaldehyde (containing 10% sucrose), and incubated for two hours using blocking solution (PBS + 10% Goat Serum + 0.2% Triton-×100; the same solution was used for the incubation with primary antibodies for 24 hours at 4°C). Cells were than incubated with the following primary antibodies: anti-CHAT (Abcam, a custom-made version of the ab181023 antibody raised in rat; diluted 1:500), anti-NUP50 (Abcam, ab137092; diluted 1:500). After overnight incubation, three washes with PBS were performed before incubating the cells with secondary antibodies (Alexa Fluor^®^ from ThermoFisher Scientific; diluted 1:1000 in PBS) for two hours at room temperature. Afterwards, cells were washed again 3 times and mounted with ProLong^TM^Gold Antifade mountant with DAPI (ThermoFisher Scientific, P36934) mixed to ibidi Mounting Medium (Ibidi, 50001).

#### Microscopy

Confocal microscopy was performed with a laser-scanning microscope (Leica DMi8) equipped with an ACS APO 63 x oil DIC immersion objective. Images were captured using the LasX software (Leica), with a resolution of 1024×1024 pixels and a number of Z-stacks (step size of 0.5 μm) enough to span the complete cell soma. To analyse the intensity of nuclear NUP50 (with ImageJ), we extracted the single plan from the complete Z-stack where nucleus of the neuron of interest was showing the largest shape.

#### Ethical approval

All procedures with human materials were in accordance with the ethical committee of Ulm University (Nr.19/12 and Nr. 135/20) and in compliance with the guidelines of the Federal Government of Germany. All participants or their next of kin gave informed consent for the study. The use of human material complied with the Declaration of Helsinki concerning Ethical Principles for Medical Research Involving Human Subjects, and experiments were performed according to the principles set out in the Department of Health and Human Services Belmont Report.

#### Mammalian cell cultures

Primary cortical neurons were prepared from C57Bl/6 mouse embryos at E18 and grown on polylysine-coated 24-well plates in neurobasal medium supplemented with 1×B27, 0.5 mM L-glutamine, and 100 IU/ml penicillin/streptomycin at 37°C with 5% CO_2_. Neurons were seeded on 6-well plates and co-transfected at day 5 using Lipofectamine 2000 (Invitrogen) with GFP plasmid plus control siRNA (Horizon # D-001810-01-20) or siRNA NUP50 (Horizon # L-042342-01-0020) at final concentration of 100 nM. Medium was replaced after 4 hours with a 1:1 (v:v) mixture of conditioned and fresh neurobasal medium. After 24 hr, determination of neuron cell death was performed by flow cytometry.

Mouse hippocampal neuronal cell HT22 were grown in Dulbecco’s Modified Eagle’s Medium (DMEM) 4.5 g/l glucose supplemented with 10% fetal calf serum and 1% penicillin– streptomycin at 37°C in 5% CO2. For immunofluorescence studies, HT22 cells were seeded 24h before transfection on glass coverslips and transfected using Lipofectamine 2000 (Fisher Scientific) for NES-mcherry-NLS reporter (Addgene plasmid # 72660, a gift from Barbara Di Ventura, Roland Eils) experiments plus siRNA control (Horizon # D-001810-01-20) or siRNA NUP50 (Horizon # L-042342-01-0020) or RNAiMAX (Fisher Scientific) with control siRNA or siRNA NUP50 at final concentration of 100 nM to analyse endogenous protein localization after NUP50 depletion.

For immunoblot analyses, HT22 cells were seeded on 6-well plates and after 24 hours, cells were transfected using RNAiMAX (Fisher Scientific) with control siRNA (Horizon # D-001810-01-20) or siRNA NUP50 (Horizon # L-042342-01-0020) at final concentration of 100 nM.

#### Immunofluorescence

For ubiquitin, RanGAP1, P62SQSTM1, Nup153, NPC, G3BP1 immunofluorescence, coverslips were incubated for 10 min in PBS with 4% paraformaldehyde, washed with PBS, and incubated in PBS plus 0.5% Triton X-100 during 10 min. The cells were washed with PBS and the coverslips were incubated during 1 h with primary antibody (1:200) against ubiquitin (3933S, Cell signaling), RanGAP1 (330800, Thermo Fisher Scientific), p62/SQSTM1 (abcam56416) Nup153 (Abcam 247000), NPC (abcam Ab24609), G3BP1 (proteintech 13057-2-AP). After washing with PBS, the coverslips were incubated with a goat anti-mouse (ThermoFisher scientific A-11001) or anti-rabbit (ThermoFisher scientific A-11008) secondary antibody conjugated with Alexa488 for 1 h, washed twice with PBS, before mounting in FluoroMount-G mounting medium with DAPI (FisherScientific # 15596276).

#### Viability assay

For cell viability, cells were detached by scraping and resuspended in PBS. TO-PRO-3 iodide (Fisher Scientific, T-3605) was added at 20 nM to each sample and gently mixed just prior to analysis, and 50,000 cells were FACS-analyzed using BD LSRII flow cytometer (BD Biosciences).

#### Western blotting

Twenty four hours after transfection, HT22 cells were collected in RIPA (TrisHCl pH8 −50mM, NaCl-150mM, EDTA pH8-1 mM, 1% Triton X-100) supplemented with protease inhibitors (Complete protease inhibitor cocktail - Merck #11697498001), incubated 30 minutes on ice and centrifuged at 4°C at 13000 rpm for 10 min. Eight µg of protein was denatured by heating at 95 °C for 10 min and loaded into 4-20% Criterion TGX stain free precasted (Biorad) and transferred onto nitrocellulose membrane using Trans-blot Turbo (BioRad). The membranes were incubated in 5% PBS-milk solution for 1 h, then with the primary antibody (1:1000) against ubiquitin (3933S, Cell signaling), RanGAP1 (330800, Thermo Fisher Scientific), p62/SQSTM1 (abcam56416) Nup153 (Abcam 247000), NPC (abcam Ab24609), G3BP1 (proteintech 13057-2-AP) in PBS overnight at 4 °C, washed 3 times with PBS and incubated with anti-mouse (Abliance BI4413) or rabbit (Abliance BI2407) horseradish peroxidase-conjugated secondary antibody. After washing three times with PBS, the membranes were developed with ECL Luminata Forte Western HRP (Millipore) and scanned with a BioRad Molecular Imager ChemiDoc XRS+. Stain free detection was used as loading control and image analyses were performed using ImageLab BioRad software.

#### Microscopy

Confocal microscopy was performed with a laser-scanning microscope (Leica DMi8) equipped with an ACS APO 63 x oil DIC immersion objective. Images were captured using the LasX software (Leica), with a resolution of 1024×1024 pixels and a number of Z-stacks (step size of 0.5 μm) enough to span the complete cell soma. To analyse the intensity of nuclear NUP50 (with ImageJ), we extracted the single plan from the complete Z-stack where nucleus of the neuron of interest was showing the largest shape.

Localisation of proteins was analysed with Widefield microscope Zeiss Imager M2 equipped with a monochrome camera Hamamatsu Orca Flash 4.0 LT.

### Drosophila experiments

#### Genetics

Flies were housed in a temperature-controlled incubator with 12:12 h on/off light cycle at 25°C. *OK371-GAL4* (Bl 26160), UAS-mCD8-GFP (Bl 5137) and *Act5C-GAL4* (Bl 4414) were obtained from the Bloomington Stock Center (Bloomington, IN). The UAS-Nup50-RNAi line was obtained from the Vienna Drosophila Resource Center (VDRC) KK library, stock number 100564. This line is predicted to have a single ON target (Nup50) and no OFF targets.

#### Quantitative real-time PCR (qPCR)

Total RNA was extracted from third instar larvae using Trizol reagent (Life Technologies). Following DNase treatment, the SensiFAST cDNA synthesis kit (BioLine) was used for reverse transcription to cDNA, using 2µg RNA as starting material. Resulting cDNA samples were used as templates for real-time PCR assays performed on a BioRad CFX system using the SensiFAST SYBR No-ROX Kit (BioLine). Primers used for quantification of Nup50 transcript levels are 5’-GTCGAGTTTAAACAGGTTGTGGAGG-3’ and 5’-GCGCGGACTAACAGTTGGATC-3’. Actin42A mRNA was used as housekeeping gene for normalization (forward primer: 5’-GCGTCGGTCAATTCAATCTT-3’ reverse primer: 5’-AAGCTGCAACCTCTTCGTCA-3’). Data were analyzed using the ΔΔCt calculation method. Experiments included no-reverse transcriptase and no-template controls.

#### Automated negative geotaxis assay to analyze motor performance

To assay motor performance, newly eclosed male flies were collected and divided into groups of 10 individuals. Until measurement, flies were maintained at 25°C with a 12-h light/dark cycle on standard *Drosophila* medium. At 1 and 7 days of age, flies were evaluated in a rapid iterative negative geotaxis assay (RING-assay), which had been previously established in the Storkebaum lab^47^. The assay is based on the innate escape response of flies to climb up the wall of a vial after being tapped down to its bottom. Flies were transferred into test tubes without anesthesia and three iterative measurements of at least 8 groups of 10 flies per genotype were video recorded with a Nikon D3100 DSLR camera. The resulting movies were converted into 8-bit grayscale TIF image sequences with 10 frames/s. Subsequently, image sequences were analyzed using an MTrack3 plug-in that automatically imports images in ImageJ, subtracts backgrounds, and filters and binarizes images to allow tracking of flies. Average climbing speed (mm/s) of all tracked flies was determined, averaged per test tube, and compared between genotypes.

#### Analysis of larval muscle innervation

To analyze synapse length on muscle 8, third instar larvae were dissected in HL3 buffer and fixed in Bouin’s for 3 min. After permeabilization and blocking (10% goat serum), immunostaining was performed with anti-Discs large 1 (anti-dlg1; DSHB, 1/200). Images were taken of muscle 8 in abdominal segment 5 using a Leica SP8 laser scanning confocal microscope with 20x Plan-Apochromat objective (0.8 NA). Maximum intensity projections of z-stacks comprising the entire NMJ were used to measure the synapse length.

#### Statistics

All data are presented as mean ± standard error of the mean (SEM) and differences were considered significant when p<0.05. Before analysis, a Robust regression and Outlier removal method (ROUT) was performed to detect statistical outliers. All data points that were considered outliers were excluded from further data analysis. For comparison of normally distributed data of two groups, unpaired t-test was used, provided that the SDs were not significantly different between groups (equal variance, evaluated by F test).

### Zebrafish experiments

#### Zebrafish Maintenance

Adult and larval zebrafish (Danio rerio) were maintained at the Imagine Institutes (Paris) fish facilities and bred according to the National and European Guidelines for Animal Welfare. Experiments were performed on wild type and transgenic embryos from AB strains as well as *Hb9:GFP* zebrafish allowing the observation of motor neurons axonal arborization within a somatic segment in fixed and live animals. Zebrafish were raised in embryo medium: 0,6 g/L aquarium salt (Instant Ocean, Blacksburg, VA) in reverse osmosis water 0,01 mg/L methylene blue. Experimental procedures were approved by the National and Institutional Ethical Committees. Embryos were staged in terms of *hours post fertilization* (hpf) based on morphological criteria (REF) and manually dechorionated using fine forceps at 24 hpf. All the experiments were conducted on morphologically normal embryos.

#### Microinjections

Morpholino antisense oligonucleotides (AMOs; GeneTools, Philomath, USA) were used to specifically knockdown the expression of the *nup50* orthologue in zebrafish (NM_201580.2) at the final concentration of 1 mM. The AUG Morpholino oligonucleotide (AMO) sequence targeting nup50 from 5’ to 3’ and complementary to the translation-blocking target is the following: GGCCATCACAGCTCAACTGAACACC, binding the mRNA target [in brackets]: cttaccagtg tgtgtgacgc gaacggttgg cgggaaac[gg tgttcagttg agctgtg(atg) gcc]aagcgga ttgcggaaaa ag. A linearized plasmid containing human NUP50 cDNA tagged by mCherry was used for the rescue experiments at the final concentration of 100 ng/µL.

#### Swimming assessment of zebrafish embryos

The locomotion of 50 hpf zebrafish embryos was measured using the *Touched-Evoked Escape Response* (TEER) test. Embryos were touched on the tail with a tip and the escape response were recorded using a Grasshopper 2 camera (Point Grey Research, Canada) at 30 frames per second. Velocity parameter was quantified per each embryo using the video tracking plugin of ImageJ 1.53 software (Sun Microsystems, USA).

#### Motor neuron morphology

50 hpf *Hb9:GFP* zebrafish embryos were fixed in 4% paraformaldehyde and captured at the same defined four somatic segments with a Spinning Disk system (Zeiss, Germany). Average axonal lengths in these segments were quantified using ImageJ 1.53 software (Sun Microsystems, USA).

## Data Availability

Annotation used in the manuscript will be made publicly available upon publication

## Acknowledgements

The authors would like to thank Andrea CHICI (Neuromuscular Disease Unit/ALS Clinic, Kantonsspital St. Gallen, St. Gallen, Switzerland), the Généthon cell and DNA bank (Evry, France) for patient DNA and lymphoblasts, and Anca MARIAN (Imagine) for technical assistance, iGenseq and iCONICS core facilities (Paris, France) for whole exome sequencing data. Image acquisition and image analysis were performed on the Imaging Platform of the CRBS, PIC-STRA UMS 38, Inserm, Unistra.

## Authors contributions

SaM and LD conceptualized the study. SaM performed statistical genetic analysis. NM, NvB and ES performed *Drosophila* experiments. SaM, JS, SD, and CS performed experiments in lymphoblasts of Patient 1, HT22 and primary neurons. AC and TB performed experiments in iPS derived motor neurons. NOA and DYH performed pathological analysis of Patient 2. AF, FM, KM, KS, JW, PMA, MW, CN, MM, AS, GL, PC, ACL, FR and StM performed exome sequencing, identified and characterized NUP50 gene variants and identified Patients 1 and 2. XM, HdC and EK performed zebrafish experiments. SDG performed histology in mouse models. KRvE and JHV performed genetic analysis and gene burden analysis. The work was supervised by SaM, StM, EK, ES, CS and LD. Manuscript was drafted by SaM and LD, and reviewed and accepted by all authors.

